# The #aware.hiv Europe study: protocol for a stepped-wedge cluster randomised trial of a multimodal hospital-based implementation strategy targeting HIV indicator condition-guided testing in European hospitals

**DOI:** 10.64898/2026.04.17.26351141

**Authors:** Klaske J. Vliegenthart-Jongbloed, Oana-Mihaela Bunea, Filip Fijołek, Isabella P. Razzolini, Tristan J. Barber, Jose I. Bernardino, Silvia Nozza, Christina K. Psomas, Marie-Angélique De Scheerder, Marta Vasylyev, Florian Voit, Carlijn C.E. Jordans, Rens Willemsen, Marianne van Wingerden, Carlo Bieńkowski, Victor D. Miron, Anna-Karina Felder, Bettina Hanssen, Jan Hontelez, Yunlei Li, Sarah E. Stutterheim, Agata Skrzat-Klapaczyńska, Oana Săndulescu, Casper Rokx, #aware.hiv Europe project

## Abstract

**Introduction:** Across Europe, many people with HIV are diagnosed late despite repeated contact with hospital services for HIV indicator conditions. These conditions flag a possible underlying HIV infection for which HIV testing is recommended. They provide an opportunity to identify people with HIV, yet implementation of indicator condition based testing remains insufficient in hospital practice. The #aware.hiv Europe study was developed to address this gap by embedding HIV teams into routine care to normalise HIV testing.

**Methods and analysis:** #aware.hiv Europe is a stepped-wedge cluster randomised trial in 30 hospitals across ten European countries. Five clusters of 6 hospitals each will sequentially transition from control to implementation periods when local HIV teams led by an infectious diseases specialist will be installed. Intervention activities include hospital-wide peer audit and feedback on missed testing opportunities, targeted education, stigma reduction activities, and strengthening of linkage to HIV prevention and care. Patients with predefined HIV indicator conditions are identified using International Classification of Diseases, 10th Revision (ICD-10) diagnosis codes, confirmed through manual review.

The primary outcome is the change in HIV testing rate among patients with confirmed HIV indicator conditions. Secondary outcomes include HIV case detection, cascades of diagnosis, care and prevention, variation in testing practices, healthcare professional knowledge and stigma, and implementation outcomes. Analyses will use mixed effects regression models accounting for clustering and time within the stepped-wedge design.

**Ethics and dissemination:** The study has ethical approval in all hospitals to use routinely collected clinical data under exemption from informed consent for patient level data. Results will be disseminated through peer reviewed publications, conferences, and collaboration with clinical and community partners with the goal to inform HIV testing policies.

**Trial registration:** ClinicalTrials.gov NCT06900829. https://clinicaltrials.gov/study/NCT06900829

**Strengths and limitations of this study:** + Large, multinational, real-world, stepped-wedge, cluster randomized trial design.

+ Primary outcome derived from routinely collected clinical data, using a GDPR- and GCP-compliant approach with exemption from informed consent.

+ Hospital-wide intervention targeting care professionals, delivered through proactive expert HIV teams across departments powered to conclude on hard HIV care cascade clinical endpoints and stigma reducing interventions.

+ Implementation science design informed by established frameworks (CFIR and RE-AIM) to strengthen cross-continental generalisability.

- Variation in healthcare systems and baseline testing practices across countries may contribute to heterogeneity in implementation and outcomes.

- Despite standardised SOPs, local clinical judgement influences the assessment of HIV indicator conditions.

## Introduction

### Background

Human immunodeficiency virus (HIV) infection is now a chronic condition. Through effective antiretroviral therapy, viral replication is stopped, preventing the development of acquired immunodeficiency syndrome (AIDS) and subsequent mortality, and preventing onward transmission. Yet across Europe, approximately half of newly diagnosed individuals were diagnosed late, reflecting prolonged periods of undiagnosed HIV infection and resulting in advanced immune damage at diagnosis. This reality has unfortunately not changed for a decade on the European continent (1, 2). Many people diagnosed with HIV had missed opportunities for testing during earlier healthcare visits, despite repeated contact with the health system (3-5). HIV indicator conditions, defined as clinical conditions associated with a higher likelihood of undiagnosed HIV infection, provide an opportunity to identify patients who should be offered testing (6). Early testing and treatment initiation avoids AIDS-related mortality, opportunistic diseases, reduces ongoing transmission, and prevents the prolonged immune activation and systemic inflammation associated with untreated HIV (7).

### Gap in current practice

International guidelines from the World Health Organization (WHO), Joint United Nations Programme on HIV/AIDS (UNAIDS), and European Centre for Disease Prevention and Control (ECDC) as well as many national guidelines consistently recommend routine HIV testing for people presenting with HIV indicator conditions (8-11). However, implementation remains highly variable and frequently inadequate in routine hospital practice. Multiple studies have demonstrated that guideline-recommended indicator condition–based HIV testing is poorly embedded in medical specialties (12-14). Systematic assessments of European clinical guidelines have shown major omissions of HIV testing recommendations for indicator conditions, contributing to low testing uptake in routine care (15). HIV stigma in healthcare settings remains substantial across Europe and may contribute to persistent gaps in HIV testing (16, 17). Earlier hospital-based implementation strategies, including electronic prompts and educational interventions, were often non-randomised, restricted to a small number of departments, and offered limited evidence on longer-term outcomes (18). There is a striking lack of large, multinational, randomised implementation trials evaluating sustainable, scalable interventions to close this diagnostic gap. In particular, there is a lack of studies that integrate clinical leadership, address behavioural change and stigma among healthcare professionals, and promote institutional testing policies to induce a hospital-wide change in testing practices.

### Rationale for the #aware.hiv Europe intervention

The #aware.hiv program was developed to address this evidence gap by embedding multidisciplinary HIV teams into routine hospital care to proactively promote, monitor, and normalize HIV testing for indicator conditions in diverse settings reflecting the cross-European clinical practice (19). Following a successful implementation pilot in the Netherlands that produced a sustained 30% absolute increase in HIV testing rates, #aware.hiv Europe aims to evaluate this strategy as a demonstration project in a multinational stepped-wedge cluster-randomized trial design across hospitals in ten European countries (20).

### Objectives

The primary objective is to determine whether implementation of HIV teams increases appropriate HIV testing among patients with HIV indicator conditions in routine care. Secondary objectives are to assess country- and condition-specific testing uptake, evaluate linkage to care and prevention, determine health-economic impacts, and explore and address stigma, as well as barriers and facilitators among healthcare professionals. The trial is thereby expected to generate robust, generalizable evidence to inform European HIV testing policy and practice in hospitals.

## Methods

### 1. Study design and setting

#### Study design

This study is a stepped-wedge cluster randomised trial, with clusters as the unit of randomisation, organised into five clusters of six hospitals each, resulting in a total of 30 participating hospitals (Figure 1: Stepped-wedge cluster randomised trial design). This design allows all clusters to implement interventions to improve HIV testing rather than remaining in the known suboptimal effective usual-care condition (as controls) for the entire trial, while the phased roll-out enables implementation with available resources.

**Figure 1:**
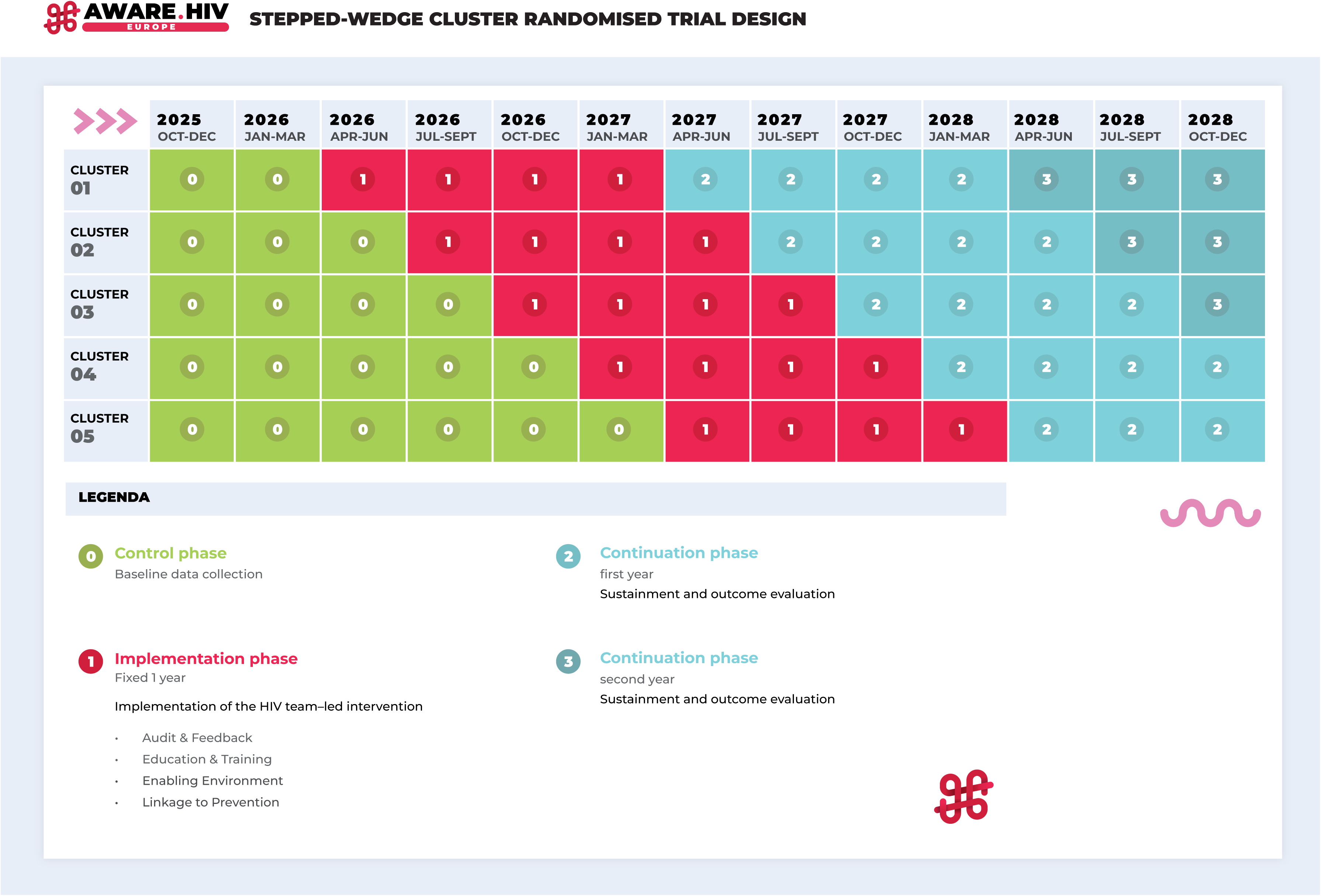
Stepped-wedge cluster randomised trial design.

Each cluster comprises six hospitals, all of which contribute data, initially as controls, with clusters then sequentially transitioning into implementation periods. Inter- and intra-cluster comparisons of outcomes over time reduce confounding and accounts for temporal trends, testing a hypothesis of interventional superiority.

#### Participating hospitals

Experienced HIV specialists (hereafter referred to country leads, CL) connected via the European AIDS Clinical Society Young Investigators Network (YING) initiated the consortium and identified eligible hospitals. These hospitals provide inpatient and/or outpatient care to adults diagnosed with HIV indicator conditions. Operational and technical capacity was assessed for feasibility, including human resources and electronic health record (EHR) infrastructure containing patient identifiers and basic clinical information. In total 31 eligible hospitals across ten European countries started local approval procedures: Belgium (1 hospital), France (1 hospital), Germany (4 hospitals), Italy (3 hospitals), the Netherlands (5 hospitals), Poland (3 hospitals), Romania (5 hospitals), Spain (3 hospitals), the United Kingdom (4 hospitals), and Ukraine (2 hospitals). Site startup was unsuccessful in Belgium where more substantial amendment requests to fit national legislation, exceeded the study timeline. Details on the 30 participating hospitals are provided in Appendix 1.

#### Patient and public involvement

Patient perspectives on HIV testing were incorporated through different levels of community engagement. General patients were consulted on HIV testing through questionnaires, and their perspectives informed the study protocol (21). The European AIDS Treatment Group (EATG) is represented partner in the study governance structure and collaborates in shaping study priorities, implementation strategies, and dissemination plans. At country level, community representatives contribute perspectives on stigma, local context, communication, and alignment with community needs. Their level of involvement varies across countries and hospital settings, ranging from consultation to active involvement in educational activities, advocacy, and linking hospital teams with community initiatives related to testing, prevention, and peer support. In addition, a community representative affiliated with Erasmus MC has been a project member since the start and co-designed the #aware.hiv artwork.

### 2. Participants

The intervention is designed to support healthcare professionals (physicians) involved in the care of adults presenting with HIV indicator conditions. Settings will include the inpatient department (IPD), outpatient department or day treatments (OPD), emergency department (EMD) and intensive care unit (ICU). The protocol enables identifying patients who have in-person contact with these healthcare professionals, and who have a newly recorded HIV indicator condition diagnosis by International Classification of Diseases, 10th Revision (ICD-10) diagnosis codes or other approved identification methods (e.g. specific laboratory results or manual clinical review). Consultations without direct patient contact or for people with known HIV infection linked to care are excluded. Any study-related contact occurs only with the treating healthcare professional.

### 3. Study procedures - HIV indicator conditions

Patients should meet the predefined criteria for an HIV indicator condition as specified in Appendix 2. Candidate indicator conditions were derived from the EuroTEST guideline and assessed based on (a) clinical relevance, (b) expected unmet testing need, (c) strength of the evidence supporting a generalised testing recommendation and (d) measurability and feasibility of reliable identification in clinical practice (6). Selection was based on independent review by country leads, followed by consensus discussions.

A predefined automated query is used to flag records containing ICD-10 codes corresponding to HIV indicator conditions. In this context, flagging refers to the automatic identification of eligible cases within the EHR, enabling their extraction.

Selection of ICD-10 codes was performed by coordinating investigators of the #aware.hiv and #aware.hiv Europe studies, based on performance of codes in the Rotterdam #aware.hiv project. As a guiding principle, codes were prioritised when an HIV indicator condition had been confirmed in more than 10% of ICD-10–flagged cases, alongside consideration of case volume and the clinical relevance of the association with HIV (22). Codes related to sexually transmitted infections and indications for post-exposure prophylaxis (PEP) were classified as prevention opportunities, with additional codes like drug use added following expert consensus.

HIV indicator conditions identified through automated queries were subsequently confirmed through manual review by investigators at each participating hospital, to ensure consistent identification of true indicator conditions and to distinguish them from alternative causes. To optimise consistency between reviewers, investigators received training, a standard operating procedure, and a predefined list of diagnoses with corresponding inclusion and exclusion criteria. After confirmation of the HIV indicator condition through manual review, records were subsequently assessed to confirm whether HIV testing had been performed.

### 4. Outcome definitions

The HIV testing rate is defined as the proportion of patients with a manually confirmed HIV indicator condition who received an adequate HIV test. Any clinically used HIV test performed within the hospital, or elsewhere with EHR documentation, from 12 months prior to the HIV indicator condition diagnosis up to the time of record review, was considered an adequate HIV test. The timing of record review after the indicator condition diagnosis was not fixed across hospitals but was kept as short as operationally feasible, typically ranging between approximately 2 weeks and 2 months. For diagnoses that may reflect acute HIV infection, such as sexually transmitted infections or mononucleosis-like illness, the HIV test had to be performed or repeated after the diagnosis of the indicator condition.

### 5. Intervention

The intervention comprises four core domains with mandatory activities, delivered hospital-wide by local HIV teams. Core components follow four scientific domains: “Implement and sustain”, “Care”, “Stigma” and “Prevention”. They collectively aim to establish a hospital-wide enabling environment for improving HIV indicator condition-based testing, including mandatory activities within each domain:

1. **Audit and feedback**, including systematic review of flagged HIV indicator conditions and written or electronic feedback to treating physicians on missed HIV testing or prevention opportunities;
2. **Education and training**, delivered through targeted departmental teaching sessions conducted at least quarterly;
3. **Activities to reduce HIV stigma** among healthcare professionals; and
4. **Strengthening pathways for linkage to HIV prevention and care**, including feedback on missed prevention opportunities and dissemination of referral pathways.

Mandatory elements are described in Appendix 3 and visualized over time in Figure 2. Activities are implemented after cluster randomisation and continue throughout a one-year implementation period, after which the primary endpoint is assessed, and hospitals enter the continuation period. Ongoing surveillance of HIV indicator conditions and HIV testing rates is conducted throughout the study using the ICD-10–based identification and manual review procedure described above. The local HIV clinical team consists of an HIV expert physician (local principal investigator), supported by a dedicated nurse, junior doctor, or research staff involved in data collection and coordination, as specified in the study standard operating procedures.

**Figure 2:**
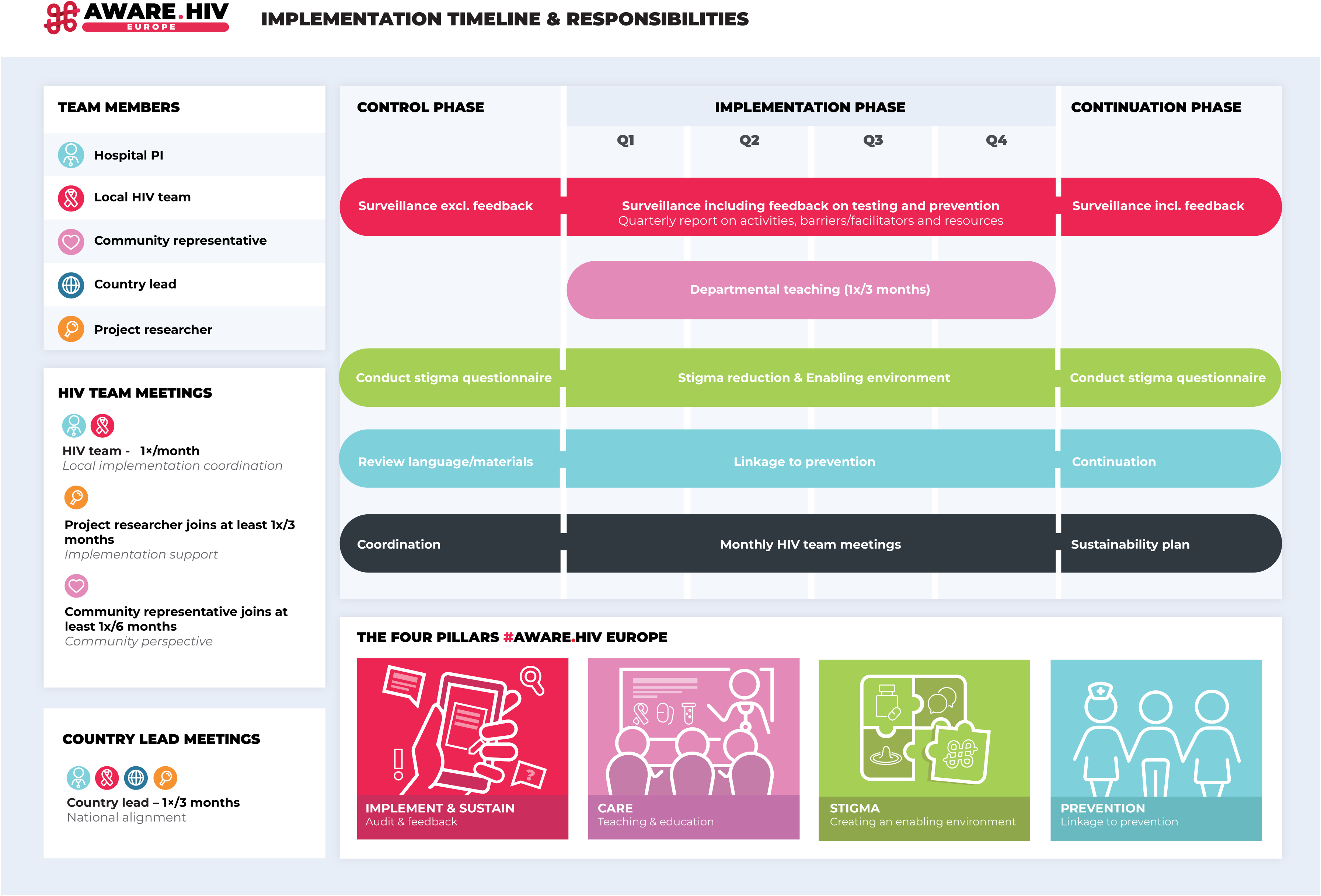
Implementation timeline and responsibilities.

Firstly, audit and feedback to treating physicians on flagged indicator conditions is a core activity. All patients receive usual care throughout the trial. Physicians are advised to offer HIV testing, but all testing and treatment decisions remain the responsibility of the treating physician in accordance with local clinical practice. Clinical decisions and testing outcomes are collected.

Secondly, the HIV team undertakes educational activities, prioritised in departments with low HIV testing rates or a high absolute number of identified missed testing opportunities, based on quarterly surveillance results. Learning objectives include improving knowledge and recognition of HIV indicator conditions, understanding the health consequences of missed HIV testing opportunities, increasing familiarity with HIV testing, prevention, and referral pathways, and raising awareness of the role of HIV-related stigma in testing practices. Pre-and post-teaching assessment of knowledge, attitude, and practice are included.

Thirdly, taking local contexts into account, we will address the ways in which stigma and discrimination impede indicator-based testing and referral pathways to HIV prevention and care. We will first measure the stereotypes, prejudice, and discrimination in health care professionals working in departments in participating health facilities, as well as structural impediments to the provision of stigma-free care. We will then address stigma-related barriers to indicator-based HIV testing and referral to HIV prevention pathways through educational activities and behaviour change interventions, and follow-up with a post-intervention survey again measuring stereotypes, prejudice, discrimination, and structural stigma to ascertain the effectiveness of the stigma reduction activities.

Fourthly, the HIV team advises on guideline recommended prevention care in identified patients with risk factors for HIV acquisition and guides treating physicians in referral pathways for patients with newly identified HIV.

These four core components are mandatory and standardised. Several elements may however be adapted by the principal investigator to the local context, including teaching formats, stigma-reduction activities, digital solutions (such as EHR prompts), and strategies to improve linkage to prevention and care, provided that mandatory activities are retained and all activities are documented in accordance with the study procedures. HIV teams receive regular feedback on HIV testing rates and missed testing opportunities to guide targeted actions. Implementation is supported by organisational structures and governance. The local HIV team meets at least monthly and coordinates activities with the country lead. Implementation activities, barriers, and HIV testing performance are reported quarterly, providing insights into performance and opportunities for improvement. Following completion of the implementation phase, hospital principal investigators and country leads develop a sustainability plan to support continuation of core intervention activities within routine care, adapted to the local context.

### 6. Implementation framework and logic model

The implementation of HIV teams for indicator condition–based testing is guided by established implementation science frameworks. The updated Consolidated Framework for Implementation Research (CFIR) is used to structure the assessment of contextual factors, barriers and facilitators, and implementation activities (23). RE-AIM is used to operationalise implementation outcomes, including reach, effectiveness, adoption and implementation(24). The continuation phase after the primary endpoint will focus on maintenance. These frameworks inform outcome selection, data collection, and the analytic strategy for implementation outcomes.

To complement these frameworks, a logic model outlines the hypothesised pathways linking implementation inputs and core intervention components to short-, intermediate-, and long-term outcomes at the healthcare professional, departmental and patient levels (Figure 3: Logic model).

**Figure 3:**
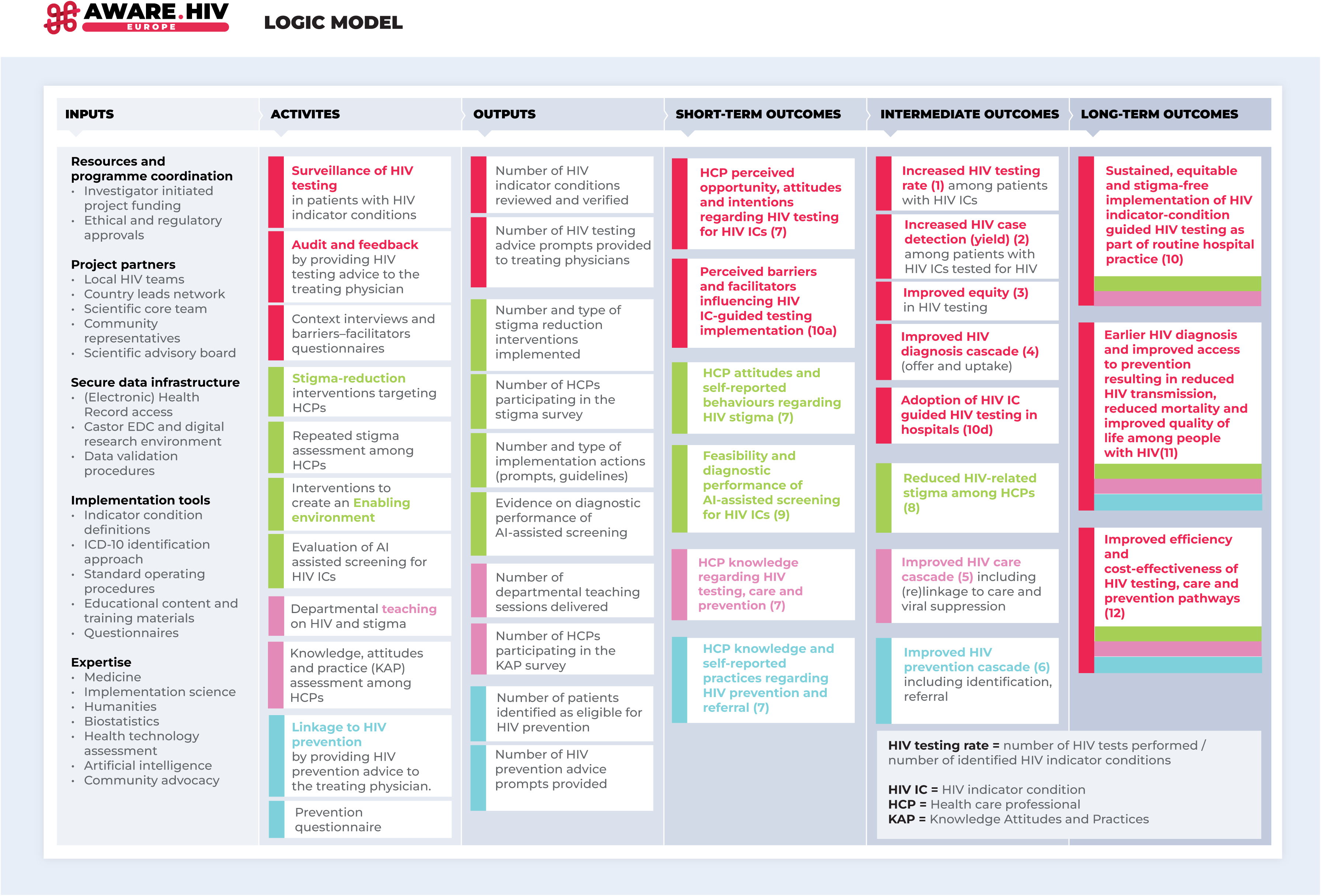
Logic model.

### 7. Outcomes

#### Primary outcome

1. The primary outcome is the change in HIV testing rate among patients with a manually confirmed HIV indicator condition. This outcome is summarised as a proportion (percentage) with 95% confidence intervals and is compared between the control phase and the one-year implementation phase per cluster within the stepped-wedge design. This outcome was selected because it directly reflects the intended behavioural change targeted by the intervention and serves as a key indicator of timely HIV diagnosis in hospital settings.

#### Secondary outcomes

Detailed definitions, measurement instruments, and analytical approaches of the secondary outcomes are provided in Appendix 4. Secondary outcomes are summarised as follows:

(2) HIV case detection, defined as the proportion (yield) as well as absolute numbers.
(3) Equity and variation in HIV testing, assessed by differences in testing rates across countries, indicator conditions, clinical specialties, time, and patient characteristics.
(4) Cascade of HIV diagnosis, describing test offer and uptake (as well as reasons for non-testing) among patients with a confirmed HIV indicator condition.
(5) Cascade of HIV care, defined as linkage or re-linkage to HIV care and clinical outcomes among newly diagnosed individuals.
(6) Cascade of HIV prevention, assessing identification of individuals eligible for HIV prevention and subsequent referral and engagement in HIV preventive services.
(7) Knowledge, attitudes and practices (KAP) among healthcare professionals, assessed using a structured questionnaire administered before and after educational activities.
(8) HIV stigma among healthcare professionals, measured using a validated self-reported stigma questionnaire.
(9) AI-assisted confirmation of HIV indicator conditions, evaluated by assessing the prediction performance, efficiency and implementation feasibility of AI-supported screening compared to manual chart review.
(10) Implementation outcomes, including contextual determinants, implementation processes, fidelity, adaptations, and outcomes assessed according to CFIR and RE-AIM frameworks.
(11) Epidemiological impact, of earlier HIV diagnoses on HIV-related morbidity, mortality, and transmission, assessed through mathematical modelling.
(12) Resource impact and cost-effectiveness, considering personnel time, implementation costs, and downstream effects such as additional HIV tests, new diagnoses, and modelled averted complications or infections.

### 7. Sample size considerations

To estimate the sample size, we conducted a power simulation study based on a Poisson mixed-effects model. The primary endpoint (HIV testing rate) will be assessed by comparing the control phase with the first year of implementation of each cluster. The study includes five clusters, each consisting of six hospitals, totalling 30 hospitals.

Data from the #aware.hiv pilot study indicate that approximately 1% of all newly diagnosed patients have a confirmed HIV indicator condition, corresponding to around 15 patients per week. The study also showed a hospital-wide increase of approximately 30% in HIV testing rates following implementation of HIV teams.

Based on #aware.hiv data and observational studies from participating countries, we assumed a mean baseline HIV testing rate of 40% and considered an absolute increase of at least 10% to be clinically relevant. Assuming a baseline HIV testing rate of 40% and a two-sided significance level of 5%, a simulation study (1,000 replications) estimated a power above 80% to detect an increase to at least 50% in the primary endpoint. Under a conservative scenario in which a participating hospital evaluates five patients with HIV indicator conditions per week, a minimum sample of 15 hospitals would be required to achieve this statistical power.

Beyond the minimum required sample size, we aimed to recruit a larger number of participating hospitals to maximise the impact of the project and support sustainable improvements in HIV testing practices. Broader participation also enables more detailed analyses of implementation processes, contextual factors, and variation between hospitals.

### 8. Randomisation and blinding

Cluster allocation and order are generated by the study biostatistician using a computerised random number generator. Clusters were balanced using restricted randomisation, stratified by country and by hospitals where the country lead also served as the local principal investigator.

The resulting allocation was concealed in sealed envelopes and was known only to the biostatistician, with cluster allocation and order disclosed to participating hospitals and investigators within three months prior to the start of the implementation phase for each cluster.

Blinding of healthcare professionals or HIV teams is not feasible given the nature of the intervention. Outcome assessment is based on routinely collected EHR data using standardised data extraction and validation procedures. To minimise bias, data collection is restricted to predefined, objective variables and follows standardised procedures as specified in the study Standard Operating Procedure (SOP) and standardised electronic Case Report Form (eCRF). Within-site comparisons, subgroup and sensitivity analysis account for possible confounders and effect modifiers.

### 9. Data collection and management

#### Data collection methods

Patient-level data on HIV indicator conditions and HIV testing are extracted from routine clinical records and entered into the central electronic data capture system (Castor EDC) by the HIV team by SOPs (Appendix 5 and 6).

During the implementation phase, local principal investigators additionally report on HIV team activities, time and resource use, as well as perceived facilitators and barriers to implementation, using structured quarterly eCRF (Appendix 7). Key barriers and facilitators were identified based on a literature systematic review on contextual elements and ranking by country leads. The online data capture tool, Castor, will be used to support stigma questionnaires in different languages. The stigma questionnaires comprise contextually relevant adaptations to theoretically-grounded and previously validated scales for measuring stigma and related constructs (REF ECDC; HPP; Earnshaw) (Appendix 8). In addition, KAP are assessed using questionnaires administered around departmental teaching sessions (Appendix 9).

For the inventory of HIV testing (Appendix 10) and inventory of HIV prevention practices (Appendix 11) at site level, a structured custom-made form is used to be completed by principal investigators. All data collection instruments are included in the protocol appendices.

#### Data quality monitoring

Data quality is promoted through standardised training of local HIV teams by data monitors and the use of eCRFs as indicated. Front-end validation at the point of data entry includes predefined answer categories, conditional logic, and validation rules, generating warnings when entries are unlikely or blocking data entry when predefined criteria for inclusion are not met. Quarterly back-end site-level data quality checks are performed by the project manager, focusing on completeness, internal consistency, plausibility, and temporal patterns of case numbers and flagged HIV indicator conditions. Data quality reports are shared with principal investigators for verification and correction, who hold final responsibility for data accuracy.

Using a risk-based remote monitoring approach, PhD researchers conduct quarterly virtual check-in meetings with participating hospitals to discuss study progress, adherence to indicator condition definitions, agreed HIV team activities, completeness and timeliness of data collection, and any practical issues encountered. In addition, the Investigator Site File (ISF) is reviewed annually in compliance with regulatory requirements, including the availability of the correct and up-to-date SOPs.

Source data remain under the responsibility of the participating hospitals and are not transferred outside the local hospitals. Pseudonymised study data are transferred to secure storage within the Sponsor (Erasmus MC) Digital Research Environment (DRE), in accordance with applicable data protection regulations and regulatory requirements. Access to the DRE is restricted to authorised members of the international scientific core team on a role basis. Access to the database is role-based and restricted to authorised study personnel. All procedures described above are specified in the Data Management Plan and Data Validation Plan and are available upon request.

A formal Data Monitoring Committee (DMC) is not established, given the implementation-focused design of the intervention, which represents recommended care, uses routinely collected data, and involves no direct patient-related interaction and study-related harms. No interim analyses before the primary endpoint or stopping rules are planned. The study biostatistician is consulted as needed in case of unexpected trends or data quality concerns.

### 10. Statistical analysis

All analyses will follow an intention-to-treat approach according to the stepped-wedge cluster randomised design. The primary analysis evaluates changes in HIV testing rates among patients with a confirmed HIV indicator condition before and after implementation of HIV teams.

The primary endpoint will be analysed using a Poisson mixed-effects regression model accounting for differences in the number of eligible patients. The model will include fixed effects for calendar time (time since study initiation) and time since intervention initiation, and random effects for cluster and hospital to account for secular trends and clustering within hospitals. Before–after comparisons will be complemented by visualisation of quarterly HIV testing rates to illustrate temporal trends throughout the study period.

Secondary outcomes will be analysed using appropriate generalised linear mixed-effects models depending on the outcome distribution. These analyses include HIV case detection, cascades of HIV diagnosis, care and prevention, equity and variation in HIV testing, and changes in healthcare professionals’ knowledge, attitudes and practices and HIV-related stigma. Detailed outcome definitions and analytical approaches are provided in Appendix 4.

Heterogeneity of intervention effects will be explored through pre-specified subgroup analyses across countries, HIV indicator conditions, clinical specialties, time, and patient characteristics. Where appropriate, interaction terms will be included in the mixed-effects models to examine variation in intervention effects across these subgroups.

If a participating site ceases data collection, it will not contribute data to the relevant analyses for the affected study period. To address under- or overperforming sites in a standardised manner, quarterly monitoring of review volume, completeness, and timeliness will take place, with targeted support where needed. Persistent deviations will be addressed in per-protocol and sensitivity analyses.

Pre-specified sensitivity analyses include:

I. exclusion of the first implementation quarter (wash-in period);
II. restriction to the first eligible presentation per patient to correct for individuals who may have multiple entries in the database;
III. exclusion of hospitals with insufficient review performance, defined as low data; completeness (<80%), inability to review the per-protocol minimum of 80 cases per quarter, or substantial delays (over 3 months) in manual review of HIV indicator conditions;
IV. exclusion of hospitals with extreme testing performance, defined as hospitals within the highest and lowest deciles of HIV testing rates or case volume;
V. exclusion of hospitals unable to review the full set of per-protocol defined HIV indicator conditions (modified per-protocol analysis).

## Ethics and exemption from informed consent

The study was approved by the Medical Ethics Review Committee of Erasmus MC, Rotterdam, the Netherlands (MEC-2024-0236; approved 8 April 2024; protocol version 1.6). Subsequent approval was obtained from the relevant ethics committees or institutional review boards of all participating hospitals, in accordance with local requirements. Substantive protocol amendments, if any, will be submitted to the relevant local research ethics committees or institutional review boards as indicated. Protocol amendments and study progress are registered on ClinicalTrials.gov (NCT06900829).

The study is conducted under an exemption from informed consent for patient-level data, as it uses routinely collected clinical data and involves no study-specific procedures at the individual patient level. This approach is justified by the large-scale and retrospective nature of the study, the impracticability of obtaining individual consent, and the risk of selection bias and unnecessary burden, including potential distress given the sensitive nature of HIV. Healthcare professionals participating in the stigma questionnaire and the KAP questionnaire provide informed consent prior to participation.

All data shared within the study are pseudonymised. Personal identifiers remain at the local site and are not transferred. Data shared with funders are aggregated only. Data handling follows the GDPR principles of data minimisation and purpose limitation, and complies with applicable data protection regulations. Secure data transfer, role-based access control, and restricted data access are applied throughout the study.

## Dissemination plan

Trial results will primarily be disseminated to healthcare professionals, participating hospitals, and the scientific community through open-access peer-reviewed publications, conference presentations, and consortium communications. In addition, the study website (www.awarehiv.com) and researchers’ professional online profiles and their network will be used to support professional dissemination. Study results will also be communicated to the wider HIV community through collaboration with patient advocacy and community organisations, supported by plain-language summaries, the study’s website and online social media accounts. Additionally, symposia will be organized which feature #aware.hiv (like the Romania advanced HIV forum). Authorship will follow the ICMJE criteria, and will reflect substantive contributions to study design, conduct, analysis, and reporting.

## Trial status

The #aware.hiv Europe study started on 1 October 2025. The first cluster randomised to start the intervention and transition to the implementation phase is planned on 1 April 2026. Completion of data collection for the primary endpoint is expected on 1 April 2028, with full data collection, including the continuation phase, expected to be completed by 1 January 2029.

## Supporting information

Appendix 4: Outcomes definitions and analyses in #aware.hiv Europe

Appendix 2: HIV indicator conditions list #aware.hiv Europe

## Author contributions

KJV-J, O-MB, FF and IPR: Conceptualization, Methodology, Writing – original draft, Writing – review & editing.

TJB, JIB, SN, CKP, M-ADS, MV, FV, CCEJ and RW: Conceptualization, Methodology, Writing – review & editing.

CB, VDM, MvW, YL, A-KF and BH: Methodology, Writing – review & editing.

SES, ASK, OS and CR: Conceptualization, Methodology, Supervision, Writing – review & editing.

CR, KJV-J: Funding acquisition.

All authors critically reviewed the manuscript, approved the final version, and agreed to be accountable for all aspects of the work.

## Transparency statement

Artificial intelligence tools were used to assist in checking completeness against the SPIRIT checklist and to support language editing of the manuscript. All content decisions and interpretations remain the responsibility of the authors.

## Data availability statement

Individual participant-level data will not be shared publicly. Requests for data access, including by consortium partners or external academic parties, are reviewed by the sponsor on a case-by-case basis, guided by the principles of purpose limitation, role-based access, and data minimisation, and subject to applicable data transfer agreements.

## Appendices

Appendix 1: Participating hospitals in #aware.hiv Europe

Appendix 2: HIV indicator conditions list #aware.hiv Europe

Appendix 3: SOP Core elements of the HIV team intervention

Appendix 4: Outcomes and definitions in #aware.hiv Europe

Appendix 5: Data dictionary #aware.hiv Europe

Appendix 6: SOP How to fill in the eCRF in Castor

Appendix 7: Questionnaire on barriers and facilitators

Appendix 8 : Stigma questionnaire

Appendix 9: Knowledge, attitudes, practices (KAP) questionnaire

Appendix 10: Documentation of local HIV testing practices and surveillance

Appendix 11: Documentation of HIV testing prevention

---

**Supplementary file 1:** Completed SPIRIT 2025 checklist for #aware.hiv Europe

