## Appendix 4: Outcomes definitions and analyses in #aware.hiv Europe for "The #aware.hiv Europe study: protocol for a stepped-wedge cluster randomised trial of a multimodal hospital-based implementation strategy targeting HIV indicator condition-guided testing in European hospitals"

| HIV indicator conditions: list and determination criteria | |
| --- | --- |
| 01 | Anal cancer * |
| 02 | Candida, esophageal |
| 03 | Candida, oral |
| 04 | Cerebral toxoplasmosis (including ocular toxoplasmosis) |
| 05 | Cervical cancer |
| 06 | Cryptococcosis, extra-pulmonary |
| 07 | Cytomegalovirus retinitis |
| 08 | Diarrhea due to cryptosporidiosis or isosporiasis more than 1 month |
| 09 | Guillain–Barré syndrome |
| 10 | Hepatitis A |
| 11 | Hepatitis B (acute or chronic) |
| 12 | Hepatitis C (acute or chronic) |
| 13 | Herpes zoster |
| 14 | Histoplasmosis |
| 15 | Invasive pneumococcal disease |
| 16 | Kaposi’s sarcoma (KS) |
| 17 | Lymphoma, Hodgkin |
| 18 | Lymphoma, non-Hodgkin |
| 19 | Mononucleosis-like illness |
| 20 | Mpox |
| 21 | Mycobacterium, other than tuberculosis * |
| 22 | Peripheral neuropathy |
| 23 | Pneumocystis carinii pneumonia (PJP) |
| 24 | Pneumonia, community-acquired (CAP) |
| 25 | Post-exposure prophylaxis (PEP) or increased risk |
| 26 | Pregnancy *(optional depending on the context, not integral to the primary endpoint)* |
| 27 | Psoriasis, severe or atypical |
| 28 | Salmonella septicemia * |
| 29 | Seborrheic dermatitis |
| 30 | Sexually transmitted infections (STI) |
| 31 | Tuberculosis (pulmonary or extrapulmonary) |
| 32 | Unexplained chronic diarrhea |
| 33 | Unexplained fever |
| 34 | Unexplained leukocytopenia/thrombocytopenia of at least 4 weeks including Idiopathic/Thrombotic thrombocytopenic purpura (ITP/TTP) * |
| 35 | Unexplained lymphadenopathy |
| 36 | Unexplained weight loss |
| 37 | HIV prevention indicators |

**General comments:**
HIV indicator conditions are selected from the online brochure “HIV Indicator Conditions: Guidance for Implementing HIV Testing in Adults in Health Care Settings”. Selection is based on expert opinion, considering prevalence, diagnostic feasibility, and clinical relevance. For feasibility, some indicator conditions have been simplified or combined (indicated with an asterisk *).
HIV indicator conditions will be identified through digitally available information.
Unless otherwise indicated, clinical diagnosis or clinical working diagnosis (without laboratory confirmation) is enough to count as indicator condition. By holding the Control (Ctrl) key and clicking on a diagnosis on the left, you will be automatically directed to the relevant page containing the inclusion and exclusion criteria.

**Multiple HIV indicator conditions:**
If multiple HIV indicator conditions are present at the same time, please select the one that has the strongest known association with HIV, using the ranking table found at the end of this document.

**Immunosuppression as an exclusion criteria**
A known immunosuppressed state—such as primary immune deficiencies, autoimmune diseases, hematological malignancies, solid organ transplantation, or treatment with immunosuppressive medication—no longer serves as a general exclusion criterion for HIV indicator conditions. In line with EuroTEST guidance, all patients presenting with an HIV indicator condition should still be considered eligible for HIV testing, regardless of these comorbidities.

The specific inclusion and exclusion criteria defined for individual indicator conditions remain in effect, as outlined in the table.

| HIV indicator conditions | Inclusion | Exclusion |
| --- | --- | --- |
| Anal cancer | Anal carcinoma, regardless of histological type (usually squamous cell carcinoma) and regardless of size or staging. | Carcinoma in situ, Bowen disease,  High-grade Squamous Intraepithelial Lesion (HSIL),  Anal Intraepithelial Neoplasia II-III (AIN II-III)  Rectum or sigmoid carcinoma |
| Candida, esophageal | The presence of *Candida* in the esophagus.  Diagnosed macroscopically (thrush) by endoscopy i.e. Oesophago-Gastro-Duodenoscopie (OGD).  Either confirmed by yeast culture or not.  If oral candida only 🡪 refer to Candida, oral | Esophageal *Candida* explained by:  - Stenosis or ulcer in esophagus.  - Radiotherapy or resection of the esophagus  Note: Immunosuppressive conditions (e.g. corticosteroids or long term antibiotics) are not considered exclusion criteria. |
| Candida, oral | Clinical diagnosis, confirmation by yeast culture is not necessary.  The presence of *Candida* in the mouth  Diagnosed macroscopically (thrush) by the eye.  If esophageal candidiasis 🡪 refer to Candida, esophageal | Note: Immunosuppressive conditions (e.g. corticosteroids or long term antibiotics) are not considered exclusion criteria. |
| Cerebral toxoplasmosis (including ocular toxoplasmosis) * | Cerebral toxoplasmosis  Based on either  * Biopsy OR * Positive PCR (Polymerase Chain reaction on CSF (Cerebrospinal Fluid) OR * Toxoplasmosis of the brain based on judgement of a radiologist, neurologist or internist/infectiologist based on the CT or MRI of the brain (multiple ring-enhancing lesions in different parts of the brain)  OR  * Toxoplasmosis in the eye (retina) based on clinical judgement of eye doctor  OR  * Serologic test for *Toxoplasma gondii* (IgM/IgG antibodies)  COMBINED WITH  * Clinical symptoms (headache, confusion, seizures, neurological deficit, altered mental status) | Presence of a positive serologic test for *Toxoplasma gondii* (IgM/IgG antibodies) without clinical or radiological suspicion |
| Cervical cancer | Based on histopathological analysis of cervical tissue (biopsy)  Independent of HPV | Cervical smear results only are excluded:  PAP 0 (not enough cells)  PAP 1 (good result)  PAP 2 (possibly self-limiting) PAP3a (mild-moderate abnormalities)  Cervix dysplasia  CIN I, II or III (Cervical Intraepithelial Neoplasia) |
| Cryptococcosis, extra-pulmonary | Clinical and laboratory evidence of extrapulmonary cryptococcosis, including but not limited to:  Cryptococcal infection of the central nervous system (CNS), skin, bone, or lymph nodes.  Confirmed by either a positive Cryptococcus antigen test OR a positive culture from extrapulmonary specimens. | Pulmonary cryptococcosis without evidence of extrapulmonary involvement.  Lack of clinical or laboratory evidence supporting the diagnosis of extrapulmonary cryptococcosis. |
| Cytomegalovirus retinitis | Clinical evidence of CMV retinitis, including but not limited to:  Presence of characteristic retinal lesions (e.g., "pizza pie" appearance, cotton-wool spots, hemorrhages).  Visual symptoms such as floaters, blurred vision, or loss of peripheral vision.  AND  Confirmation of CMV retinitis diagnosis by ophthalmologic examination, including indirect ophthalmoscopy and fundoscopic evaluation. | Lack of clinical or ophthalmologic evidence supporting the diagnosis of CMV retinitis.  Presence of another ocular condition or retinal pathology that could mimic symptoms of CMV retinitis (e.g., retinal detachment, ocular toxoplasmosis).  If ocular toxoplasmosis 🡪 Cerebral toxoplasmosis |
| Diarrhea due to cryptosporidiosis or isosporiasis > 1 month | Intestinal infection by the protozoan parasites *Cryptosporidium* spp. or *Isospora belli.*  Diagnosed by either: * Stool examination with staining (oocysts) * PCR (DNA) * Immunofluorescence of stool (antigens) * EIA (enzyme immunoessay) for antigen detection.  The duration should be > 1 month.  If chronic diarrhea, but no cryptosporidiosis or isosporiasis 🡪 refer to ‘unexplained chronic diarrhea’ | Note: Immunosuppressive conditions (e.g. inflammatory bowel disease (IBD) like Crohn’s or ulcerative colitis) are not considered exclusion criteria.   Note: Guillain-Barré syndrome (GBS) is often preceded and triggered by trauma, toxins, or an infection, but these should *not* be considered exclusion criteria:  - Campylobacter jejuni,  - Cytomegalovirus (CMV),  - Epstein-Barr virus (EBV),  - Mycoplasma pneumoniae,  - Vaccination (influenza)  - Varicella-zoster virus (chickenpox and shingles),  - Surgery, medical procedures, trauma, and stress. |
| Guillain–Barré syndrome | Clinical diagnosis by a neurologist or other specialist doctor. Usually based on symptoms such as symmetric muscle weakness, starting in the lower extremities with ascending paralysis, loss of reflexes, paresthesias, and autonomic dysfunction. Sometimes supported by CSF (Cerebrospinal Fluid) analysis, EMG (electromyography), blood tests or imaging. |  |
| Hepatitis A (acute) | Acute hepatitis A only.  Diagnosed with either one of the following tests: * Hepatitis A RNA testing (PCR) of the blood * Hepatitis A IgM Antibody Test | Hepatitis A IgG Antibody Test alone, as this is indicating past infection or vaccination. |
| Hepatitis B (acute or chronic) | Regardless of duration or Regardless of disease activity.  Diagnosed with any test. | Presence of Hepatitis B Surface Antibody (anti-HBs), regardless of vaccination state.  Diagnosis solely based on imaging (ultrasound or FibroScan) or liver tests (such as alanine aminotransferase (ALT) and aspartate aminotransferase (AST)), with no virological proof. |
| Hepatitis C (acute or chronic) | Regardless of duration, chronicity, and disease activity  Diagnosed with either one of the following tests:  * HCV Antibody Test (Anti-HCV)  * HCV RNA Test (PCR) | Presence of anti-HCV, but registered treatment for HCV (hepatitis C virus) in the past.  Diagnosis solely based on imaging (ultrasound or FibroScan) or liver tests (such as alanine aminotransferase (ALT) and aspartate aminotransferase (AST)), with no virological prove. |
| Herpes zoster (=shingles, reactivation of varicella zoster virus) | Clinical diagnosis by any doctor.  Characterized by a painful, unilateral rash that usually appears as a band or strip following a specific dermatome of fluid-filled blisters.  Virological confirmation (PCR, viral culture or varicella zoster antibodies) is not necessary.  All patients, regardless of age.  Also, if presenting with postherpetic neuralgy or secondary infection. If HIV is not ruled out earlier during the same disease period.  Regardless of location (trunk, face, eye area, scalp). | Primo infection (varicella zoster, chicken pox)  Note: Immunosuppressive conditions (e.g. autoimmune disease, transplantation, or immunosuppressive medication) are not considered exclusion criteria.  Note: Age is not considered an exclusion criterion. |
| Histoplasmosis | Clinical and radiographic evidence of histoplasmosis resulting in treatment:  * Fever, cough, fatigue, and shortness of breath.  * Pulmonary infiltrates or mediastinal lymphadenopathy  With or without laboratory confirmation:  * Histopathological examination demonstrating intracellular yeast forms or positive antigen testing for *Histoplasma capsulatum*. | Note: Immunosuppressive conditions (e.g. autoimmune disease, transplantation, or immunosuppressive medication) are not considered exclusion criteria. |
| Invasive pneumococcal disease | This includes but is not restricted to meningitis, bacteremia, septic arthritis, osteomyelitis, pneumonia, endocarditis.  Diagnosed by identifying: pneumococci in  extra pulmonary material  OR  Pneumococci in sterile material OR  Positive pneumococcal antigen test in urine | Pneumococci from the upper airways  Otitis media Sinusitis Conjunctivitis  Bronchitis  If pneumonia, but no proven invasive disease 🡪 assess the criteria for community acquired pneumonia |
| Kaposi’s sarcoma (KS) | Clinical or histopathological evidence of Kaposi sarcoma, including but not limited to:  Presence of characteristic skin lesions (purple, red, or brown patches, plaques, or nodules).  Involvement of mucosal surfaces, lymph nodes, or internal organs (e.g., gastrointestinal tract, lungs).  Confirmation of Kaposi sarcoma diagnosis by biopsy is not necessary to establish KS as an indicator condition. | Presence of an alternative etiology for skin lesions or mucosal abnormalities, such as other types of malignancy (e.g., melanoma, squamous cell carcinoma) or non-neoplastic skin conditions (e.g., dermatitis, psoriasis).  If HHV-8–related lymphoma → refer to the section ‘Lymphoma, non-Hodgkin’ |
| Lymphoma, Hodgkin | Hodgkin lymphoma |  |
| Lymphoma, non-Hodgkin | ***B-cell lymphoid proliferations and non-Hodgkin lymphomas*** | |
|  | Strong connection to HIV: **Large B-cell lymphomas**, including subtypes, especially:  • Diffuse Large B cell lymphoma (DLBCL)  • EBV-positive diffuse large B-cell lymphoma  • Plasmablastic lymphoma (PBL)  • Primary central nervous system lymphoma (PCNSL), also known as Primary large B-cell lymphoma of immune-privileged sites  **Burkitt lymphoma/leukemia (BL)**  **KSHV/HHV8-associated B-cell lymphoid proliferations** and lymphomas, especially:  • Primary effusion lymphoma (PEL)  • Extracavitary PEL (EC-PEL)  • KSHV/HHV8-associated multicentric Castleman disease (MCD)  • KSHV/HHV8-positive diffuse large B-cell lymphoma (KSHV/HHV8-DLBCL)  Poor connection to HIV: **Precursor B-cell neoplasms**: B-cell lymphoblastic leukaemias (B-ALL) or lymphomas (B-LBL)  **Pre-neoplastic and neoplastic small lymphocytic proliferations**  e.g. Monoclonal B-cell lymphocytosis (MBL), Chronic lymphocytic leukaemia (CLL), small lymphocytic lymphoma (SLL)  **Splenic B-cell lymphomas and leukaemias** (e.g. Hairy cell leukaemia, Splenic marginal zone lymphoma)  **Indolent B-cel lymphoma’s e.g.** Lymphoplasmacytic lymphoma (LPL)  Marginal zone lymphoma (MZL) including MALT lymphoma  Follicular lymphoma (FL)  Mantle cell lymphoma (MCL)  **Tumour like lesions with B-cell predominance other than KSHV/HHV-8 associated Castleman disease**  Unicentric Castleman disease  Idiopathic multicentric Castleman disease  IgG-related disease | **Lymphoid proliferations and lymphomas associated with immune deficiency and dysregulation** Post-transplant lymphoproliferative disorder (PTLD)  Lymphoid proliferations and lymphomas associated with a primary (inborn) immune deficiency Lymphomas related to congenital syndromes  Plasma cell neoplasms and other diseases with paraproteins e.g. cold agglutinin disease, monoclonal gammopathy of undetermined significance (MGUS), amyloidosis, heavy chain disease, plasmacytoma or POEMS syndrome.  If unexplained lymphadenopathy 🡪 refer to the section ‘unexplained lymphadenopathy’  If mycobacterial infection 🡪 refer to the section ‘tuberculosis’  🡪or refer to the section ‘mycobacterial infection other than tuberculosis’ |
|  | ***T-cell and NK-cell lymphoid proliferations and lymphomas*** | |
|  | **Tumour-like lesions with T-cell predominance**  **Precursus T-cell neoplasms** like T-lymphoblastic leukaemia (T-ALL) /lymphoma (T-LBL)  **Mature T-cell and NK cell leukaemia’s**, for example:  • T-prolymphocytic leukemia  • Adult T-cell leukaemia or lymphoma (ATLL)  • Sézary syndrome  • Extranodal NK/T-cellymphoma, nasal type  • EBV-positive NK/T-cell lymphomas  **Primary cutaneous T-cell lymphoma’s** including Mycosis fungoides, especially  • Peripheral T-cell lymphoma, not otherwise specified  • HTLV related Primary cutaneous T-cell lymphomas  **Intestinal or hepatosplenic T-cell and NK-cell lymphoid proliferations and lymphomas**  **Anaplastic large cell lymphoma**  **Nodal T-follicular helper (TFH) cell lymphoma**, formally known as Angioimmunoblastic T-cell lymphoma (AITL) or Follicular T-cell lymphoma |  |
| Mononucleosis-like illness | Acute tonsillitis with negative EBV (Epstein-Barr Virus) test  * negative EBV serology OR  * negative EBV PCR  Especially if: * nontender lymphadenopathy * macular or maculopapular exanthem generalizing from face, chest to extremities – including palms and soles * duration > 1 week * insufficient response on antibiotics | Acute tonsillitis for which no additional testing was done  Negative EBV serology or PCR, performed because of hematological/ immunological disease or related to solid organ transplantation.  Other explanation of tonsillitis: EBV (Epstein-Barr Virus), CMV (cytomegalovirus), HHV-6 (human herpesvirus 6), HSV-1 (herpes simplex virus), group A beta-hemolytic *Streptococcus*, adenovirus  Dengue  Chikungunya  Chronic tonsillitis  If unexplained fever or a possible acute retroviral illness 🡪 refer to the section ‘unexplained fever’ |
| Mpox (or monkeypox) | Clinical diagnosis suffices if consistent with monkeypox, characterized by the presence of a rash exhibiting papules, vesicles, and crust formation.  Laboratory confirmation via PCR (polymerase chain reaction) or serological tests is not obligatory. | Other explanation for similar clinical symptoms, such as chickenpox, herpes zoster, or other viral skin infections.  If herpes zoster 🡪 refer to the section ‘herpes zoster’ |
| Mycobacterium, other than tuberculosis (disseminated or extrapulmonary) | Disseminated (widespread spread of an infection from its original site to multiple organs or body systems)  OR extrapulmonary (infections occurring outside the lungs)  Including but not restricted to: *Mycobacterium avium* complex (MAC), *M. kansasii, M. xenopi, M. xenopi, M. chelonae, M. fortuitum,*  *M. abscessus* | If the diagnosis is classical tuberculosis caused by *M. tuberculosis* 🡪 refer to the category “tuberculosis” |
| Peripheral neuropathy | Unexplained polyneuropathy: symmetrical  and feet>hands  Clinical diagnosis by a medical doctor.  Characteristic profile (not mandatory): * quickly progressing * painful * confirmed with EMG (electromyogram) * Reduced vibrating and cold sensation  * Reduced ankles stretch reflex | If the neuropathy is explained by: - Pressure neuropathy - Polyneuropathy in diabetes mellitus or alcoholism - Postoperative neuropathic pain  - Neuropathy of nervus ulnaris  - Iatrogenic nerve injury - Traumatic nerve injury - Due to B12 deficiency, nitrous oxide, MGUS, ACNES or neuroma - Hypo/hyperthyroidism |
| Pneumocystis carinii pneumonia (PJP or PCP) | Diagnosed by a specialist doctor.  Characteristic profile (not mandatory):  * Fever and chills * Dry cough * Dyspnea, out of proportion to the physical exam finding * Chest pain * Bilateral interstitial infiltrates with ground-glass appearance * Raised LDH (lactate dehydrogenase) or beta-D-Glucan in the blood * Microbiological confirmation on BAL (bronchoalveolar lavage) or sputum with immunofluorescence staining or PCR (DNA) | Note: Immunosuppressive conditions (e.g. autoimmune disease, transplantation, or immunosuppressive medication) are not considered exclusion criteria. |
| Pneumonia, community-acquired | Any pneumonia requiring treatment, either based on clinical judgement or radiological imaging.  If a specific causative agent, refer to the respective indicator conditions  🡪 “invasive pneumococcal disease”  🡪 “Pneumocystis jirovecii pneumonia” 🡪 “Tuberculosis” | Sarcoidosis  Asbestosis  Influenza A, COVID-19 or viral RTI  Interstitial pneumonia due to GIST (Gastrointestinal Stromal Tumor)  Aspiration pneumonia Obstructive pneumonia  Cryptogenic organizing pneumonia (COP)  Hospital acquired pneumonia (onset after ≥ 48 h of hospital admission or within 3 months of a previous admission)  Non-infectious pneumonitis  Note: Immunosuppressive conditions (e.g. autoimmune disease, transplantation, or immunosuppressive medication) are not considered exclusion criteria. |
| Post-exposure prophylaxis or increased risk for contracting HIV | Individuals receiving post-exposure prophylaxis (PEP) for potential HIV exposure.  Individuals identified as being at increased risk for contracting HIV (e.g., based on behavioral risk factors or belonging to key groups) | Absence of post-exposure prophylaxis (PEP) treatment.  Individuals not identified as being at increased risk for contracting HIV.  Post-exposure prophylaxis for indications other than HIV, such as meningococci or rabies.   - Note: Individuals identified as having an increased risk based on partner notification are recorded under 'partner notification' in the 'Baseline characteristics' section rather than under 'HIV Indicator condition'. |
| Pregnancy | Pregnancy confirmed through diagnostic testing or ultrasound.  Irrespective of pregnancy trimester, complications, or outcome. | Pregnancy not confirmed by diagnostic testing or ultrasound, relying solely on medical history. |
| Psoriasis, severe or atypical | A new, fast progressing psoriasis  Psoriasis with erythrodermia (red skin)  Psoriasis with reactive arthritis  Combination of seborrheic eczema and psoriasis (sebopsoriasis)  Impetiginized psoriasis  Different phenotypes of psoriasis combined | Easy to treat, typical psoriasis |
| Salmonella septicemia* | Presence of *Salmonella* bacteria in the bloodstream.  Diagnosed by blood culture or PCR (DNA) on blood or bone marrow.  Including but not restricted to:  *S.* Typhi *S.* Paratyphi Non-typhoidal salmonella (NTS) including *S. enteritidis* and *S. typhimurium*. | *Salmonella* infection which is restricted to the gastrointestinal tract.  A diagnosis based only on a serological test (Enzyme-linked immunosorbent assay, ELISA). |
| Seborrheic dermatitis | Extensive or difficult to treat seborrheic dermatitis Impetiginized seborrheic dermatitis (wet areas with yellow exudate, crustae or signs of inflammation)  Generalized erythroderma | Atopic eczema  Acrovesiculous/dyshidrotic eczema  Eczema hyperkeratoticum  Asteatotic eczema  Hypostatic eczema  Eczema of vulva/perineal area Patients referred for patch test for allergy in eczema. |
| Sexually transmitted infections (STI) | All STIs, including:  - syndromically approached/treated  - Pelvic inflammatory disease (PID) - Condylomata accuminata - Scabies with crustae (*Sarcoptes scabiei*)  Other STIs: Chlamydia including Lymphogranuloma Venereum (LGV) (*Chlamydia trachomatis*)  Gonorrhea (*Neisseria gonorrhoeae*)  Syphilis, lues (*Treponema pallidum*)  Herpes simplex virus, cold sores (HSV)  Human papillomavirus (HPV)  Trichomoniasis (*Trichomonas vaginalis*)  *Mycoplasma genitalium* | Uncomplicated scabies *Ureaplasma* spp.  Bacterial Vaginosis (BV)  If mpox🡪 refer to the section ‘Mpox’ |
| Tuberculosis (pulmonary or extrapulmonary) | Diagnosed by a doctor.  Requiring TB treatment.  “Active TB”  Either with positive or negative sputum. Either in the lungs or elsewhere, like pleuritis, lymphadenitis, osteoarticular, meningitis, abdominal, pericardial, genitourinary.  Characteristic profile (not mandatory):  * Persistent cough (>3 weeks), hemoptysis, weight loss, night sweats.  * X-ray with infiltrates or cavities. * Sputum with *M. tuberculosis* based on mycobacterial culture, microscopy (acid-fast bacilli AFB in Ziehl-Neelsen, Kiyoun or Auramine staining) or PCR (e.g. GeneXpert)  Active TB (also if treatment initiated but sputum negative) | Latent TB.  Positive Tuberculin Skin Test (TST) like Mantoux or Interferon-Gamma Release Assays (IGRAs) like QuantiFERON with no signs of active disease.  Treatment of a latent TB usually involves one or two drugs (isoniazid INH, rifampin RIF or rifapentine PRT) as opposed to four drugs for an active TB.  If the diagnosis caused by a tuberculosis species, other than *M. tuberculosis* 🡪 refer to the category “Mycobacterial infection, other than tuberculosis”  Note: Immunosuppressive conditions (e.g. autoimmune disease, transplantation, or immunosuppressive medication) are not considered exclusion criteria. |
| Unexplained chronic diarrhea | If ≥ 3 liquid bowel movements per day AND > 4 consecutive weeks without interruption.  HIV should be part of the initial workup.  If: Cryptosporidiosis or  Isosporiasis 🡪 refer to “Diarrhea due to cryptosporidiosis or isosporiasis”  If one of the following etiologies is confirmed, still count as HIV indicator condition:  Microsporidiosis e.g., *Enterocytozoon bieneusi* CMV colitis *Mycobacterium avium* complex (MAC)  Salmonellosis  Campylobacteriosis  Shigellosis | Another etiology known for chronic diarrhea, like:  - Inflammatory bowel disease (IBD), e.g. Crohn’s disease, ulcerative colitis - Malabsorption syndrome, e.g. celiac disease, lactose intolerance, pancreatic insufficiency/chronic pancreatitis  - Infections: *Giardia lamblia*, *Entamoeba histolytica*)  - Microscopic colitis, e.g. collagenous colitis, lymphocytic colitis - Hyperthyroidism, carcinoid tumors and neuroendocrine tumors, Zollinger-Ellison syndrome, Verner-Morrison syndrome - Ischemic colitis - Graft-versus-host-disease (GVHD) |
| Unexplained fever | No known etiology or working hypothesis.  Analysis of fever with no other explanation.  OR  Acute retroviral syndrome (ARS), characterized by  * fever  AND * lymphadenopathy AND * maculopapular rash (often also: malaise, headache, pain behind the eyes, muscle pain, throat pain, diarrhea, peripheral neuropathy) Incubation time of 2-4 weeks. | Fever with another explanation |
| Unexplained leukocytopenia or thrombocytopenia  lasting at least 4 weeks (including Idiopathic/thrombotic thrombocytopenic purpura (ITP/TTP)) * | Leukocytopenia or thrombocytopenia with a duration of > 4 weeks with no other explanation.  Cut off: Leukocytes < 4*10^9/L Thrombocytes < 150 *10^9/L  OR  Clinical diagnosis of ITP  OR  Clinical diagnosis of TTP  Characteristic profile of ITP (not mandatory):  * Low platelets (<100) * Easy bruising or petechiae * Antiplatelet antibodies  Characteristic profile of TTP (not mandatory): * Low platelets * thrombotic microangiopathy (small clots)  * hemolytic anemia with schistocytes in peripheral blood smear * neurologic symptoms * renal impairment * fever/malaise  * relatively normal PT and aPTT  * reduced ADAMTS13 activity | Leukocytopenia or thrombocytopenia with a duration of < 4 weeks  Leukocytopenia or thrombocytopenia with another explanation.  Other explanation like:  - Bone marrow disorder, e.g. aplastic anemia, myelodysplastic syndromes (MDS), leukemia, myelofibrosis - Autoimmune disorder e.g. systemic lupus erythematosus (SLE), rheumatoid arthritis (RA) - Medication e.g. chemotherapy drugs, immunosuppressive medications - Vitamin deficiency e.g. folate or B12 deficiency  - Connective tissue disorders e.g.  Sjögren's syndrome or Behçet's disease - Hypersplenism  - Disseminated intravascular coagulation (DIC) or sepsis |
| Unexplained lymphadenopathy | Enlarged lymph nodes with no other explanation.  If lymphoma or Castleman disease 🡪 refer to the section ‘Lymphoma’  If tuberculosis 🡪 refer to the section ‘Tuberculosis’  If mycobacterial infection, other than tuberculosis 🡪 refer to the section ‘Mycobacterial infection, other than tuberculosis’ | Other explanation like:  - Infections e.g. streptococcal, staphylococcal, cat scratch fever (*Bartonella henselae)*, EBV (Epstein-Barr virus), CMV (cytomegalovirus) or toxoplasmosis.  - Autoimmune disorder e.g. systemic lupus erythematosus (SLE), rheumatoid arthritis (RA)  - Metastatic cancer - Inflammatory disease e.g. sarcoidosis, Kawasaki disease or Familial Mediterranean Fever (FMF)  - Hemophagocytic lymphohistiocytosis (HLH) |
| Unexplained weight loss | Referral for weight loss with no other explanation.  If tuberculosis 🡪refer to the section ‘Tuberculosis’ | Other explanation like: - Cancer - Inflammatory bowel disease (IBD) e.g. Crohn’s disease or ulcerative colitis  - Poorly controlled diabetes, hyperthyroidism - Severe chronic obstructive pulmonary disease (COPD) or congestive heart failure (CHF)  - Psychiatric or neurological disorders, e.g. dementia, Parkinson’s disease, depression, eating disorder like anorexia nervosa, substance abuse - Malabsorption syndrome, e.g. celiac disease, lactose intolerance, pancreatic insufficiency/chronic pancreatitis Exclude if a weight loss less than 5% weight in 6 months is documented. |

**Ranking table: Multiple HIV indicator conditions at the same time**

If multiple HIV-related conditions are concurrent, please prioritize identifying the condition most strongly linked with HIV. Note that AIDS-defining illnesses carry a stronger association with HIV compared to non-AIDS-defining illnesses. The higher the ranking, the stronger the connection with HIV.

**AIDS-defining Illnesses**

Pneumocystis carinii pneumonia (PJP)

Kaposi’s sarcoma (KS)

Cerebral toxoplasmosis (including ocular toxoplasmosis)

Cytomegalovirus retinitis

Cryptococcosis, extra-pulmonary

Lymphoma, non-Hodgkin

Histoplasmosis

Candida, esophageal

Tuberculosis (pulmonary or extrapulmonary)

Mycobacterium, other than tuberculosis

Cervical cancer

Diarrhea due to cryptosporidiosis or isosporiasis > 1 month

Salmonella septicemia

**Non-AIDS-defining Illnesses**

Hepatitis C (acute or chronic)

Hepatitis B (acute or chronic)

Mononucleosis-like illness

Sexually transmitted infections (STI)

Invasive pneumococcal disease

Unexplained chronic leukocytopenia/thrombocytopenia/ITP/TTP

Unexplained fever

Herpes zoster

Lymphoma, Hodgkin

Seborrheic dermatitis

Mpox

Anal cancer

Post-exposure prophylaxis (PEP) or increased risk

Pneumonia, community-acquired (CAP)

Unexplained chronic diarrhea

Unexplained lymphadenopathy

Unexplained weight loss

Hepatitis A

Candida, oral

Psoriasis, severe or atypical

Guillain–Barré syndrome

Peripheral neuropathy

Pregnancy
