## Appendix 2: HIV indicator conditions list #aware.hiv Europe for "The #aware.hiv Europe study: protocol for a stepped-wedge cluster randomised trial of a multimodal hospital-based implementation strategy targeting HIV indicator condition-guided testing in European hospitals"

**INTRODUCTION**

This appendix provides the detailed specification of all primary and secondary study outcomes, in accordance with the SPIRIT recommendations. For each outcome, the operational definition, data source, measurement approach, and planned analysis are described. This appendix complements the outcome overview presented in the main manuscript and supports methodological transparency and reproducibility across study sites.

**PRIMARY OUTCOME**

**(1) HIV testing rate**

**Outcome definition:** Change in HIV testing among patients with a confirmed HIV indicator condition following implementation of HIV teams.

**Variable:** HIV testing rate, defined as the proportion of patients with a manually confirmed HIV indicator condition who had a documented HIV test within the window from 1 year prior to the HIV indicator condition diagnosis up to the time of record review. A valid HIV test is defined as any clinically used HIV test (laboratory-based or point-of-care) recorded in the electronic patient record.
For HIV indicator conditions reflecting possible acute HIV infection, including sexually transmitted infections or mononucleosis-like illness, the HIV test must have been performed or repeated after the diagnosis was made.

**Summary measure:** Proportion (%) with 95% confidence intervals.

**Time point:** Control phase versus 1-year implementation phase within each cluster (stepped-wedge design).

**Analysis:** The primary analysis will use a Poisson mixed-effects regression model, accounting for differences in the number of eligible patients, including time since study initiation and time since intervention initiation as fixed effects, and cluster and hospital as random effects, to account for secular trends and within-hospital correlation. Analyses will follow an intention-to-treat approach, whereby hospitals are analysed according to their assigned intervention timing irrespective of the degree of implementation.

**Rationale:** This outcome was selected as it directly reflects the primary behavioural change targeted by the intervention and serves as a key indicator of timely HIV diagnosis in hospital settings.

**SECONDARY OUTCOMES**

**(2) HIV case detection, defined as the proportion (yield) as well as absolute numbers**

**Outcome definition:** Change in HIV case detection among patients presenting with a confirmed HIV indicator condition following implementation of HIV teams, assessed both as the proportion of positive tests (yield) and as the absolute number of newly diagnosed HIV infections.

**Variables:
2a. HIV test yield**
Defined as the proportion of newly diagnosed HIV infections among patients with a confirmed HIV indicator condition who underwent HIV testing.

**2b. Absolute HIV case detection**
Defined as the number of newly diagnosed HIV infections among patients presenting with a confirmed HIV indicator condition within each hospital and study period.

**Summary measure:** HIV test yield will be summarised as proportions (%) with 95% confidence intervals. Absolute HIV case detection will be summarised as counts of newly diagnosed HIV infections per hospital and study period.

**Time point:** Across the study period, comparing the control phase and implementation phase.

**Analysis:** Mixed-effects regression models will be used to assess changes in HIV test yield over time, accounting for intervention status, time since study initiation, and clustering by hospital and cluster. Absolute HIV case detection will be analysed using mixed-effects count models accounting for time, intervention status, and clustering by hospital and cluster. Exploratory analyses will assess HIV case detection among patients tested following documented audit and feedback activities

**Rationale:** HIV test yield reflects the diagnostic efficiency of targeted HIV testing among patients with HIV indicator conditions and enables comparison across hospitals independent of patient volume. Absolute case detection reflects the population-level impact of the intervention in terms of additional HIV diagnoses identified through implementation of HIV teams.

**(3) Equity and variation in HIV testing, assessed by differences in testing rates across countries, indicator conditions, clinical specialties, time, and patient characteristics.**

**Outcome definition:** Equity and variation in HIV testing rates among patients with a confirmed HIV indicator condition across clinical, geographical and temporal contexts.

**Variable:** HIV testing rate, defined as the proportion of patients with a confirmed HIV indicator condition who received a documented HIV test.

**Outcome measures:** Variation in HIV testing rates will be assessed across the following predefined dimensions:
 3a. variation within countries
 3b. variation between countries
 3c. variation across HIV indicator conditions
 3d. variation across medical specialties
 3e. variation over time
 3f. variation by patient characteristics (age and sex)

**Summary measure:** Proportions (%) with comparative estimates across subgroups.

**Time point:** Across the study period, comparing control and implementation phases.

**Analysis:** Variation in HIV testing rates will be analysed using generalised linear mixed-effects models with Poisson distribution, including intervention status and subgroup variables as fixed effects, and hospital and cluster as random effects. Patient characteristics will be summarised descriptively using counts and proportions for categorical variables and means (SD) or medians (IQR) for continuous variables, as appropriate.

**Rationale:** This outcome evaluates the robustness and equity of the intervention across different clinical, geographical and temporal settings and explores whether HIV indicator condition–guided testing is implemented consistently across patient groups and care contexts.

**(4) Cascade of HIV diagnosis, describing test offer and uptake (as well as reasons for non-testing) among patients with confirmed HIV indicator conditions.**

**Outcome definition:** The cascade of HIV diagnosis describes the number of people with a confirmed HIV indicator condition, subsequent HIV test offer and acceptance, as well as expressed barriers to testing.

**Variable:** Cascade of HIV diagnosis, defined as sequential steps among patients with a confirmed HIV indicator condition.

**Outcome measures**
 4a. Focus population: number of patients with a confirmed HIV indicator condition
 4b. Testing offer (reach): proportion of patients for whom an HIV test was offered
 4c. Testing acceptance (uptake): proportion of patients who accepted the HIV test, operationalised as completion of HIV testing among those offered testing. Where testing was not performed, acceptance or refusal will be inferred from documented reasons for non-testing
 4d. Consent procedures: documentation of consent procedures where applicable
 4e. Reasons for not testing: documented reasons for not offering or not performing HIV testing

**Summary measure:** Counts and proportions at each step of the cascade, with the focus population set at 100%.

**Time point:** Across the study period, comparing the control and implementation phase.

**Analysis:** The cascade of HIV diagnosis will be presented descriptively using counts and proportions. For step-specific analyses, denominators will be defined conditionally on completion of the preceding cascade step (i.e. testing offer among patients with a confirmed HIV indicator condition, and testing acceptance among patients to whom an HIV test was offered). Variation in cascade outcomes will be assessed within and between countries, across HIV indicator conditions, across medical specialties, and over time. Where appropriate, logistic regression or mixed-effects models will be used to analyse the probability of testing offer and testing acceptance, including relevant covariates and accounting for clustering at hospital and cluster level. Where applicable, testing offer and acceptance will be derived documented testing and recorded reasons for non-testing, including patient refusal.

**Rationale:** This outcome identifies missed opportunities for HIV testing along the diagnostic pathway and provides insight into barriers affecting testing offer and uptake, thereby informing targeted implementation and quality improvement strategies.

**(5) Cascade of HIV care, defined as linkage or re-linkage to HIV care and clinical outcomes among newly diagnosed individuals.**

**Outcome definition:** The cascade of HIV care describes the number of people with a newly diagnosed HIV infection who are linked to HIV care services following presentation with an HIV indicator condition.

**Variable:** Cascade of HIV care among patients newly diagnosed with HIV following presentation with an HIV indicator condition.

**Outcome measures**
 5a. Focus population: number of patients with a confirmed HIV indicator condition and a subsequent HIV diagnosis
 5b. (Re)linkage to routine HIV care (reach): proportion of newly diagnosed patients linked or re-linked to HIV care services
 5c. Viral suppression (uptake): proportion of patients achieving viral suppression according to routine clinical data

**Summary measure:** Counts and proportions at each step of the cascade of care.

**Time point:** Across the study period, comparing the control and implementation phase.

**Analysis:** The cascade of HIV care will be described using counts and proportions. For step-specific analyses, denominators will be defined conditionally on completion of the preceding cascade step (i.e. linkage to care among newly diagnosed patients, and viral suppression among patients linked to care). Variations in linkage to care and viral suppression will be assessed within and between countries, across HIV indicator conditions, across specialties, and over time. Multivariable regression or mixed-effects models will be used where appropriate to explore determinants of successful linkage to care and viral suppression, accounting for clustering by hospital and cluster. Where linkage to care or viral suppression is not observed, documented reasons will be described descriptively where available,

**Rationale:** This outcome evaluates whether improved HIV testing leads to timely linkage to care and effective treatment outcomes, thereby assessing the downstream clinical impact of the intervention beyond diagnosis alone.

**(6) Cascade of HIV prevention, assessing identification, referral, and engagement in HIV prevention services among eligible patients.**

**Variable:** Cascade of HIV prevention among patients presenting with a confirmed HIV indicator condition, defined as the following sequential steps.

**Outcome measures:**
6a. Focus population: number of patients with a confirmed HIV indicator condition who meet predefined eligibility criteria for HIV prevention
6b. Appropriate referral (reach): proportion of eligible patients for whom a referral or documented advice for HIV preventive services
6c. Uptake for prevention (uptake): proportion of referred patients who initiated or engaged with HIV preventive services, where such information is available in routine clinical records
6d. Variation in the cascade of HIV prevention: assessed within and between countries, across HIV indicator conditions, across medical specialties, and over time, taking into account the local and national availability of HIV preventive services and access characteristics (Appendix 11)

**Summary measure:** Counts and proportions at each step of the cascade, with the focus population set at 100%. Absolute and relative differences in cascade outcomes between control and implementation phases.

**Time point:** Across the study period, comparing control and implementation phases.

**Analysis:** The cascade of HIV prevention will be described using counts and proportions. For step-specific analyses, denominators will be defined conditionally on completion of the preceding cascade step (i.e. referral to prevention among eligible patients, and uptake of prevention among patients who received a referral or documented advice). Variations in referral and uptake for prevention will be analysed using logistic or generalised linear mixed-effects models, as appropriate, including fixed effects for time and intervention status and random effects for hospital and cluster to account for clustering and temporal trends. Where appropriate, multivariable models will be used to explore factors associated with referral to and uptake of HIV preventive services, including healthcare professional characteristics, stigma and knowledge, attitudes and practices measures, and contextual implementation factors.

**Rationale:** This outcome evaluates whether improved HIV testing and case finding through HIV teams translates into timely identification of individuals eligible for HIV prevention and effective linkage to preventive services, thereby addressing missed prevention opportunities.

**(7) Knowledge, attitudes and practices (KAP) among healthcare professionals, assessed using a structured questionnaire administered before and after educational activities.**

**Outcome definition:**  The level and change of knowledge, attitudes and practices regarding HIV testing, prevention and care among healthcare professionals.

**Descriptive statistics:** Descriptive statistics will include absolute and relative frequencies (n/N, %) for categorical variables and mean with standard deviation (SD) for continuous variables. Median and interquartile range (IQR) will be reported for continuous variables with skewed distribution. The data distribution will be assessed prior to further analyses.

**Main analysis approach:** Individual ordinal Likert-scale items will be combined into composite scores for each domain of the questionnaire (i.e., Knowledge, Attitude, and Practice). These scores will be treated as continuous variables, as they are derived from multiple items, and they are assumed to approximate a normal distribution.

**Handling of clustering and planned subgroup analyses:** Subsequent analyses will adopt a paired approach, as the same participants are assessed before and after the educational intervention. Differences between pre- and post-intervention scores will be evaluated using a paired-samples t-test for variables with a normal distribution. For variables that do not meet normality assumptions, non-parametric tests will be applied. Subgroup analyses will be conducted to assess changes in scores within predefined groups, including country, department, job title, duration of professional experience, and age. Departments will be grouped in three groups: surgical departments, acute departments, and medical departments. Occupation will be divided into four groups: medical specialist, resident, nurse, and other.

**Approach to handling missing data:** All primary analyses will be conducted on a per-protocol basis, including only participants who completed both pre- and post-intervention assessments. Sensitivity analyses may be performed using an intention-to-treat approach, if appropriate. The proportion of missing responses will be reported for each variable and questionnaire domain. The potential impact of missing data on the study results will be evaluated and discussed.

**Psychometric properties:** The psychometric properties of the questionnaire will be evaluated by assessing internal consistency using Cronbach’s alpha coefficients for each domain (Knowledge, Attitude, and Practice), calculated separately for pre- and post-intervention data. To ensure relevance and representativeness, the questionnaire was reviewed by member of Scientific Core and Country Leads of #aware.hiv Europe.

**Outcome measure 7a. KAP change following departmental education**

**Variable:** Change in KAP domain scores (knowledge, attitudes, practices), measured using a KAP questionnaire (Appendix 9) administered before and after departmental educational sessions.

**Summary measure:** For each KAP domain, scores will be summarised as mean (SD). Where appropriate, median (IQR) will be reported descriptively to reflect the ordinal nature of the underlying Likert items.

**Time point:** Before versus after departmental educational sessions.

**Analysis:** Changes in KAP domain scores will be analysed using linear mixed-effects models, treating domain scores as continuous outcomes, and accounting for repeated measurements and clustering within hospitals.

**Outcome measure 7b. Association between KAP and study endpoints**

**Variable:** Association between KAP domain scores (knowledge, attitudes, practices; level and change) and the primary and secondary study endpoints, including HIV testing rate, new HIV diagnoses, and cascades of HIV diagnosis, care and prevention.

**Summary measure:** Rate ratios (RR) with 95% confidence intervals for rate-based outcomes
Odds ratios (OR) with 95% confidence intervals for binary cascade outcomes

**Time point:** Across the study period, aligned with the timing of KAP assessments and outcome assessment.

**Analysis:** Associations between KAP domain scores (baseline levels and pre–post changes) and study endpoints will be analysed using generalised linear mixed-effects models, with appropriate link functions depending on the outcome. Models will include fixed effects for time and intervention status, and random effects for cluster and hospital, to account for temporal trends and clustering.

**Rationale:** This secondary outcome evaluates whether departmental education improves healthcare professionals’ HIV-related knowledge, attitudes and practices, and whether KAP levels and changes are associated with the state of or improvements in HIV testing behaviour and downstream cascades of diagnosis, care and prevention.

**(8) HIV stigma among healthcare professionals, measured using a validated self-reported stigma questionnaire.**

**Variable:** HIV stigma among healthcare professionals, measured using a structured stigma questionnaire (Appendix 8).

**Outcome measures:**
8a. Fear of HIV infection: proportion of healthcare professionals expressing fear of occupational HIV transmission
8b. Institutional-level facilitators or barriers: proportion reporting institutional factors that facilitate or hinder HIV testing and care
8c. Negative attitudes towards people with HIV: proportion expressing stigmatizing attitudes
8d. Observed stigma: proportion reporting observed stigmatizing behaviours or practices in the clinical setting

**Summary measure:** Proportions (percentages) with 95% confidence intervals.

**Time point:** Before and after implementation of the HIV team intervention, including baseline and follow-up assessments during the study period.

**Analysis:** Changes in stigma indicators over time will be analysed using logistic mixed-effects models, accounting for repeated measurements and clustering within hospitals. Variation in HIV stigma indicators will be assessed within and between countries, across HIV indicator conditions, across medical specialties, and over time.

**Rationale:** This outcome evaluates the prevalence and determinants of HIV-related stigma among healthcare professionals and assesses whether implementation of HIV teams is associated with reductions in stigma.

**(9) Effectiveness of AI-assisted HIV indicator condition confirmation**

**Variable:** Use and impact of AI-assisted pre-screening to support identification of HIV indicator conditions in electronic health records, based on predefined ICD-10 codes and explicit inclusion and exclusion criteria, compared with gold-standard manual chart review.

**Outcome measures:**
9a. AI-assisted case identification efficiency: Proportion of charts requiring manual review with AI-assisted pre-screening compared with standard manual screening without AI support.
9b. Prediction performance of AI-assisted case finding: Sensitivity, specificity, positive predictive value, and negative predictive value of AI-assisted chart screening, using manual chart review decisions as the ground truth.
9c. Mean or median reviewer time spent per chart, for non-AI-assisted screening, as well as total reviewer time across a predefined review period. To assess how many time can be saved using AI. Time estimates may additionally be stratified or adjusted according to chart classification outcome (e.g., included vs excluded), where relevant.
9d. Implementation factors: Acceptability, appropriateness, and feasibility of AI-assisted screening as perceived by HIV team members, clinicians, and other relevant reviewers involved in chart assessment.
9e. Decision transparency and reasoning: Proportion of AI-screened charts for which the system provides explicit and reviewable reasoning for inclusion or exclusion based on the predefined criteria, and proportion of these AI-generated decisions judged by human reviewers to be clinically interpretable and sufficiently justified.

**Summary measure:**
Proportions (percentages), medians with interquartile ranges, with 95% confidence intervals where appropriate.

**Analysis**:
AI-assisted chart screening will be evaluated using a retrospective validation approach. All charts included in the analysis will first be identified using preselected ICD-10 codes corresponding to HIV indicator conditions, consistent with the standard case identification strategy used in the #aware.hiv project. These charts are then manual reviewed by the HIV team, whose decisions serve as the ground truth based on the predefined inclusion and exclusion criteria.
The AI-assisted screening tool will be retrospectively applied to the same set of charts as a pre-screening tool in future intended use. The original manual chart review will be completed first and will remain unchanged. AI-generated screening decisions (include/exclude) will then be compared with the decisions of the completed manual chart review. This primary evaluation assesses retrospective AI-supported pre-screening rather than real-time collaborative decision-making between AI and human reviewers.
Diagnostic performance metrics will be calculated, including sensitivity, specificity, positive predictive value, and negative predictive value of AI-assisted screening relative to manual review. A 2×2 classification table will be constructed to quantify true positives, false positives, true negatives, and false negatives.

Efficiency outcomes will be assessed by estimating the proportion of charts requiring manual review under a retrospective AI-assisted pre-screening scenario compared with full manual review. Reviewer time per chart and total reviewer time across a defined review period will be measured in a separate evaluation, which may compare chart assessment with and without AI support.

Additional analyses may explore variation in AI performance and efficiency across HIV indicator conditions, clinical specialties, and participating study sites. Decision transparency will be assessed by examining whether AI-assisted screening outputs provide explicit reasoning for inclusion or exclusion based on the predefined criteria, and whether this reasoning is judged by reviewers to be clinically interpretable. Where feasible, discrepant cases between AI-assisted screening and manual review may undergo additional expert adjudication to explore whether apparent AI false positives include clinically relevant cases missed during the initial manual review.

**Rationale:**
This retrospective analysis evaluates whether AI-assisted pre-screening can support HIV teams by improving the efficiency of identifying charts with potential HIV testing opportunities while reducing manual workload. In addition, where discrepant cases are further adjudicated, the analysis may explore whether AI-assisted screening identifies clinically relevant cases missed during the initial manual review. It also assesses the feasibility, acceptability, and interpretability of integrating AI-supported screening into hospital workflows.

**(10) Implementation outcomes, including contextual determinants, implementation processes, fidelity, adaptations, and outcomes assessed according to CFIR and RE-AIM frameworks.**

**Outcome measure 10a. Contextual factors (CFIR domains I–IV): Contextual characteristics across CFIR domains I–IV.**

**Outcome definition:**
Contextual determinants influencing the implementation of HIV indicator condition–guided testing in participating hospitals, covering CFIR domains I–IV: intervention characteristics, outer setting, inner setting, and characteristics of individuals involved in implementation.

**Variable:** Contextual implementation determinants identified across the following CFIR domains:

I) Intervention characteristics (e.g. perceived complexity, adaptability, and strength of evidence)
II) Outer setting (e.g. national policies, healthcare system factors, societal attitudes towards HIV)
III) Inner setting (e.g. organisational structures, resources, communication, leadership engagement)
IV) Characteristics of individuals (e.g. knowledge, beliefs, and self-efficacy of healthcare professionals)

**Data sources and data collection:**
Contextual data will be collected using two complementary approaches:

1. Semi-structured interviews with country leads and local principal investigators prior to the start of the implementation phase, using a CFIR-based interview guide structured according to the four CFIR domains (Appendix 7).
2. Healthcare professional surveys, including the Knowledge, Attitudes and Practices (KAP) questionnaire (Appendix 9), capturing individual-level determinants such as knowledge, attitudes towards HIV testing, perceived stigma, and reported clinical practices.

**Summary measure:** Findings will be synthesised into contextual implementation profiles for each participating hospital and country, summarising key facilitators and barriers across CFIR domains. Results will be presented as:

- thematic summaries of qualitative interview findings
- descriptive summaries of KAP questionnaire results (e.g. domain scores)
- integrated contextual profiles highlighting major determinants of implementation.

**Time point:** Context assessment will primarily take place during the control phase prior to the start of implementation, with survey data providing additional insight into baseline characteristics of healthcare professionals.

**Analysis:** Qualitative interview data will be analysed using thematic analysis guided by the CFIR framework, combining deductive coding based on CFIR constructs with inductive identification of emergent themes. Coding will be performed using qualitative analysis software.

Quantitative KAP data will be analysed descriptively to characterise baseline levels of knowledge, attitudes and practices among healthcare professionals.

Findings from interviews and survey data will be integrated to develop contextual interpretations of implementation variation across hospitals and countries. Contextual determinants will be used to support interpretation of implementation outcomes, fidelity, and effectiveness results in subsequent analyses.

**Rationale:** Understanding contextual determinants is essential for interpreting variation in implementation success and anticipating facilitators and barriers across diverse healthcare systems. By combining qualitative CFIR-guided interviews with quantitative survey data on healthcare professional knowledge, attitudes and practices, this outcome provides a structured description of the organisational and individual context in which the intervention is implemented.

**Outcome measure 10b. Implementation activities, adaptations and fidelity
(CFIR domain V; RE-AIM “Implementation”)**

**Outcome definition:** Implementation activities performed by local HIV teams, including the processes through which the intervention is delivered, adaptations made to the implementation approach, and the degree of fidelity to the intended intervention components.

**Variable:**
Implementation processes executed by local HIV teams, including:

- implementation activities across key intervention components
- adaptations made to the implementation strategy
- fidelity to core intervention elements
- resources and personnel time required for implementation activities

**Data sources and data collection:**
Implementation data will be collected prospectively using a structured implementation monitoring tool (Appendix 7), completed by local principal investigators or designated HIV team members.

The tool captures:

- implementation activities across intervention domains (e.g. surveillance, audit and feedback, education, stigma reduction, enabling environment, and linkage to prevention and care)
- qualitative descriptions of adaptations to the implementation approach
- reporting of major internal or external events affecting implementation
- personnel time and resources required to conduct implementation activities

These data are used both to characterise implementation processes and to inform the economic evaluation (Outcome 12).

**Summary measure:** Implementation activities will be summarised as:

- counts and descriptive summaries of activities per hospital and reporting period
- qualitative descriptions of implementation adaptations
- descriptive summaries of implementation events affecting the intervention

Implementation fidelity will be summarised using domain-specific and overall fidelity scores.

**Time point:** Implementation data will be collected:

- every 3 months during the implementation phase
- every 6 months during the continuation phase.

**Analysis:**
Implementation fidelity will be assessed using predefined indicators grouped into three domains:

1. HIV team functioning (teaming and teaching)
2. audit and feedback activities
3. enabling environment and linkage to care and prevention

Each indicator will be scored using a three-point fidelity scale:

0 = not implemented
1 = partially implemented
2 = fully implemented

For indicators based on activity frequency (e.g. meetings or teaching sessions), numerical responses will be converted into fidelity scores based on adherence to the recommended frequency defined in the study Standard Operating Procedure.

For each domain, item-level scores will be averaged to generate a domain-specific fidelity score. An overall fidelity score will subsequently be calculated as the mean of the three domain scores, resulting in a score ranging from 0 to 2, with higher scores indicating greater adherence to the intended implementation of the intervention.

Implementation activities, fidelity scores, and reported adaptations will be summarised descriptively across hospitals and reporting periods. Patterns over time and between hospitals will be explored graphically and descriptively.

Where data allow, exploratory analyses will assess associations between implementation fidelity and implementation outcomes, including HIV testing rates and cascade indicators, using mixed-effects models accounting for clustering by hospital and time period.

**Rationale:** Documenting implementation activities, adaptations, and fidelity provides insight into how the intervention is delivered in practice across diverse hospital settings. These data support interpretation of variation in effectiveness outcomes and enable identification of implementation strategies associated with successful adoption and sustainability.

**Outcome measure 10c. Barriers and facilitators over time (CFIR-based)**

**Outcome definition:**
Perceived barriers and facilitators influencing implementation of the HIV indicator condition–guided testing intervention, assessed longitudinally during the implementation process.

**Variable:**
Implementation determinants across predefined CFIR constructs related to: innovation, outer setting, inner setting, individuals involved, and the implementation process.

**Data sources and data collection:**
Barriers and facilitators will be assessed using a standardised CFIR-based questionnaire (Appendix 7), completed by the local principal investigator or designated HIV team member responsible for implementation activities.

The questionnaire contains predefined items mapped to CFIR constructs. For each item, respondents classify the determinant as:

- barrier
- neutral factor
- facilitator

Additional open-text responses allow respondents to describe important contextual factors influencing implementation.

**Summary measure:** For each CFIR construct and questionnaire item, responses will be summarised as:

- proportions of responses classified as barrier, neutral, or facilitator
- descriptive summaries of qualitative explanations where provided

Results will be summarised per hospital and reporting period.

**Time point:**
The questionnaire will be administered:

- every 3 months during the implementation phase
- every 6 months during the continuation phase

**Analysis:**
Barriers and facilitators will be analysed descriptively over time to identify patterns in perceived implementation determinants across hospitals. For each CFIR construct, the distribution of responses (barrier, neutral, facilitator) will be summarised per hospital and reporting period. These distributions will be visualised using graphical displays, such as bar charts or stacked bar plots, to illustrate the relative proportion of barriers and facilitators across CFIR domains and over time.

Changes in the distribution of barriers and facilitators will be examined:

- over time within hospitals
- between hospitals and countries
- across CFIR domains

Where data allow, exploratory analyses will assess whether reported barriers and facilitators are associated with implementation outcomes such as implementation fidelity, implementation activities, or HIV testing outcomes, using mixed-effects models accounting for clustering by hospital and time period.

**Rationale:** Monitoring barriers and facilitators over time allows identification of determinants that hinder or support implementation during different stages of the intervention. This information supports interpretation of implementation success, guides potential adjustments to implementation strategies, and informs sustainability of the intervention across diverse hospital settings. Repeated assessment allows tracking how perceived determinants evolve during implementation.

**Outcome measure 10d. Implementation outcomes synthesised using the RE-AIM framework**

**Outcome definition:** Implementation outcomes synthesised across the RE-AIM dimensions (Reach, Effectiveness, Adoption, Implementation, and Maintenance) to provide an integrated interpretation of how the HIV team intervention is implemented and sustained across participating hospitals. **Variable:** Implementation outcomes mapped to the RE-AIM dimensions using predefined study outcomes and implementation measures.

Key dimensions include:

- Reach: The extent to which the intervention reaches the target population (patients with HIV indicator conditions and healthcare professionals involved in their care).
- Effectiveness: The impact of the intervention on key clinical and public health outcomes.
- Adoption: The extent to which hospitals and clinical departments initiate and adopt indicator condition–guided HIV testing practices.
- Implementation: The extent to which the intervention is delivered as intended, including fidelity, adaptations and implementation processes.
- Maintenance: The extent to which the intervention is sustained over time after the initial implementation phase..

**Data sources and data collection:** No additional data collection is conducted specifically for RE-AIM; instead, the framework is used to integrate existing study outcomes and implementation findings.

**Summary measure:** For each RE-AIM dimension, descriptive summaries will be generated per hospital and across the study period. Results will be presented using a combination of:

- descriptive summaries of implementation indicators
- graphical representations to visualise variation between hospitals and over time
- integrated RE-AIM summaries linking effectiveness and implementation findings.

**Time point:** RE-AIM outcomes will be synthesised across the implementation phase, with maintenance assessed during the continuation phase.

**Analysis:** RE-AIM dimensions will be analysed descriptively to provide an integrated interpretation of implementation success and sustainability.

Results from contextual analyses (Outcome 10a), implementation activities and fidelity (Outcome 10b), and barriers and facilitators over time (Outcome 10c) will be interpreted together with effectiveness outcomes to assess how implementation processes influence intervention impact across hospitals.

Visual summaries (e.g. dashboards or graphical summaries) may be used to illustrate patterns across RE-AIM dimensions and facilitate comparison between sites.

**Rationale:** The RE-AIM framework provides a structured approach to integrating effectiveness and implementation findings. By synthesising outcomes across RE-AIM dimensions, the study aims to understand not only whether the intervention works, but also how it is adopted, implemented, and sustained across diverse hospital settings.

| **RE-AIM** | **Focus** | **Key measures** | **Outcome** |
| --- | --- | --- | --- |
| **Reach** | Patients with HIV IC reached for testing, care or prevention | • Patients reached for HIV testing  • Patients reached for HIV care  • Patients reached for HIV prevention | **4**  **5**  **6** |
|  | Healthcare professionals | • Health care professionals reached by feedback • Departments reached by teaching | **3**  **7, 10b** |
| **Effectiveness** | Patients with HIV indicator conditions | • HIV testing rate  • HIV case detection (yield and absolute numbers)  • Linkage to HIV care and viral suppression  • Linkage to HIV prevention services  • Earlier HIV diagnosis and modelled health impact | **1**  **2** **5** **6** **11** |
| **Adoption** | Hospitals | • Initiation and uptake of IC-based testing  • Variation between hospitals and countries | **1** **10** |
| **Implementation** | HIV teams/ processes | • Implementation activities  • Adaptations to the implementation approach  • Resources (staff, time, tools)  • Barriers and facilitators affecting implementation | **10b, 10c** |
| **Maintenance** | Hospitals (continuation phase) | • Sustainability of testing practices  • Continuation of implementation activities (maintenance plan) | **10b** |

**(11) Epidemiological impact, assessed by evaluating earlier HIV diagnoses and, using modelling analyses, estimating effects on HIV-related morbidity, mortality, and future HIV transmission.**

**Outcome measure 11a. Expediting of HIV diagnosis: Timeliness of HIV diagnosis.**

**Variable:** CD4 T-cell count at HIV diagnosis and stage of disease at diagnosis, including CDC classification and presence of advanced immunodeficiency (CD4 T-cell count <200 cells/mm³).

**Summary measure:** Mean (SD) and IQR CD4 T-cell count at diagnosis; proportion of patients with advanced immunodeficiency (CD4 T-cell count <200 cells/mm³); and distribution of CDC stage at diagnosis.

**Time point:** Control phase versus implementation phase.

**Analysis:** Mixed-effects models accounting for time, intervention status, and clustering by hospital and cluster. The control phase serves as the primary counterfactual, reflecting the expected timing of HIV diagnosis in the absence of the intervention.
If statistical power is insufficient for within-study comparisons, CD4 T-cell counts and stage at diagnosis during the implementation phase will be compared descriptively with national surveillance data on CD4 T-cell count distribution and disease stage at HIV diagnosis, ideally derived from hospital-based populations where available.
If sufficient power is available, subgroup analyses will be conducted by country and by strata of baseline HIV testing rates across participating hospitals.

**Rationale:** Delayed HIV diagnosis remains common despite prior healthcare encounters in which HIV testing could have been offered. Indicator condition–guided testing aims to identify missed testing opportunities and advance the timing of HIV diagnosis compared with usual practice, where diagnosis may occur later in the course of infection. Because the spectrum of HIV indicator conditions includes both earlier clinical presentations and AIDS-defining illnesses, the intervention may identify individuals across a broad range of disease stages, including patients presenting with advanced disease. Timeliness of diagnosis will therefore be assessed using complementary indicators, including CD4 T-cell count, the proportion with advanced immunodeficiency (CD4 T-cell count <200 cells/mm³), and CDC stage at diagnosis.

Increased case detection may initially identify a higher proportion of patients with advanced or previously missed HIV infection, which may lower the average CD4 count at diagnosis despite improved testing performance. Therefore, markers of diagnostic timeliness will not be interpreted in isolation, but in conjunction with HIV case detection, linkage to care, and modelled downstream health impact.

**Outcome measure 11b. Modelled downstream health impact: Estimated impact of HIV diagnosis on HIV-related morbidity, mortality, and health burden (DALYs/QALYs).**

**Variable:** Modelled estimates derived from observed changes in diagnostic timeliness and HIV testing outcomes in the study.

**Summary measure:** Estimated number of averted HIV-related complications and deaths, and estimated gains in quality-adjusted life years (QALYs) or disability-adjusted life years (DALYs).

**Analysis:** Individual-based modelling will be used to simulate HIV disease progression using established relationships between CD4 T-cell count at diagnosis, disease progression, treatment initiation, and mortality. Model parameters will be informed by published literature and calibrated using observed study outcomes (including testing rates and diagnostic timeliness). Sensitivity analyses will explore uncertainty in key model parameters.

**Rationale:** Earlier HIV diagnosis enables earlier initiation of antiretroviral therapy, which reduces HIV-related morbidity and mortality. Because these long-term outcomes cannot be directly observed within the time frame of the trial, modelling approaches are used to translate empirical study outcomes into expected downstream health impacts.

**Outcome measure 11c. Modelled impact on future HIV transmission: Estimated reduction in future HIV transmission resulting from earlier HIV diagnosis and treatment.**

**Variable:** Modelled estimates of averted secondary HIV infections attributable to HIV diagnosis observed in the study.

**Summary measure:** Estimated number and relative reduction of secondary HIV infections over defined time horizons.

**Analysis:** Individual-based transmission modelling will be used to estimate the impact of earlier HIV diagnosis on onward HIV transmission. The model will incorporate established relationships between HIV diagnosis, treatment initiation, viral suppression, and infectiousness. Model parameters will be informed by published literature and calibrated to reproduce transmission dynamics and estimated HIV incidence levels in European settings. Scenario and sensitivity analyses will explore uncertainty in key assumptions.

**Rationale:** Earlier HIV diagnosis enables earlier initiation of antiretroviral therapy, which reduces viral load and infectiousness. As a result, the duration of untreated infection and the probability of transmission are reduced. Because changes in HIV incidence cannot be directly measured within the time frame of the study, modelling approaches are used to estimate the potential population-level impact of reduced transmission.

**(12) Resource impact and cost-effectiveness, considering personnel time, implementation costs, and downstream effects such as additional HIV tests, new diagnoses, and modelled health gains.**

**Outcome measure 12a. Project resource use and costs: Resource utilisation and costs per hospital.**

**Outcome:** Project-related resource utilisation and costs per hospital.

**Variables**

- Self-reported personnel time spent on the project (questionnaire based)
- Implementation-related resources (e.g. data infrastructure)
- HIV testing procedures (laboratory tests and confirmatory testing)

**Summary measures**

- Total personnel hours per hospital
- Total costs (€) per hospital per reporting period

**Time points**

- Every 3 months during the implementation phase
- Every 6 months during the continuation phase

**Analysis:** Descriptive cost analysis from a health care sector perspective. Resource use is multiplied by locally applicable unit costs (e.g. salaries, laboratory tariffs), allowing comparison across hospitals while preserving local cost structures.

**Rationale:** Previous work shows that personnel time and laboratory testing constitute the largest cost drivers of indicator condition–based HIV testing interventions. Quantifying these elements empirically provides realistic estimates of the resources required for implementation and sustainability across diverse hospital settings

**Outcome measure 12b. Cost-effectiveness: Cost-effectiveness of HIV indicator condition–based testing support.**

**Outcome:** Cost-effectiveness of HIV indicator condition–based testing support

**Variables**

- Unit cost per HIV test
- Cost per new HIV diagnosis
- Incremental cost per additional HIV test (where applicable)
- Incremental cost per additional HIV diagnosis (where applicable)
- Cost per quality adjusted life year (QALY) gained

**Summary measures**

- Absolute cost-effectiveness estimates
- Incremental cost-effectiveness estimates (pre- vs post-implementation, where feasible)

**Analysis:** Cost-effectiveness is calculated by relating total or incremental costs to observed outcomes in terms of HIV tests performed and new diagnoses identified. Analyses distinguish between:

- Empirical costs, reflecting all observed testing and implementation costs
- Direct project-related costs, capturing costs directly attributable to testing recommendations
- Hypothetical fully implemented scenarios, exploring cost-effectiveness under complete testing uptake
- Modelled cost-effectiveness expressed as cost per QALY gained, incorporating model-based estimates of downstream health impact (Outcome 11b) and transmission effects (Outcome 11c), together with empirical cost estimates and literature-derived disability weights.

**Rationale:** Empirical studies of targeted HIV testing demonstrate that cost per diagnosis as well as QALY-based modelling are robust and policy-relevant outcomes. Cost-effectiveness is strongly influenced by underlying HIV prevalence, testing uptake, and local unit costs, underscoring the importance of context-specific interpretation rather than universal thresholds
